# Bacterial genome wide association study substantiates *papGII* of *Escherichia coli* as a patient independent driver of urosepsis

**DOI:** 10.1101/2023.05.24.23290482

**Authors:** Aline Cuénod, Jessica Agnetti, Helena Seth-Smith, Tim Roloff, Denise Wälchli, Dimitri Scherbakov, Rashid Akbergenov, Sarah Tschudin-Sutter, Stefano Bassetti, Martin Siegemund, Christian H. Nickel, Jacob Moran-Gilad, Timothy G. Keys, Valentin Pflüger, Nicholas R. Thomson, Adrian Egli

## Abstract

Urinary tract infections are extremely common and often caused by *Escherichia coli*. Bacterial virulence factors and patient characteristics have been linked separately to progressive, invasive infection. The interaction of these factors has however rarely been considered. We whole genome sequenced 1076 *E. coli* isolates recovered from urine-or blood culture samples from 825 clinical cases. The majority of isolates belonged to the phylogroups B2 and D and encoded polysaccharide capsules. In line with previous studies, our bGWAS identified *papGII* to be associated with bacteraemia. In a generalised linear model correcting for patient characteristics, *papGII* was substantiated as a major contributor to invasive infection. Further, an independent cohort of 1,657 urine samples was PCR screened for *papGII* carrying *E. coli*, confirming the increased relative frequency of *papGII*+ strains to cause invasive infection. This study builds on previous work linking *papGII* with invasive infection by showing that it is a patient-independent risk factor that has diagnostic potential.

## Introduction

Urinary Tract Infections (UTIs) are among the most common diseases, affecting >150 million people each year worldwide ^1^. Up to 60% of women suffer from at least one symptomatic UTI in their lifetime, with ∼10% of women experiencing a symptomatic UTI every year ^2^. *Escherichia coli* is the most common cause of UTI ^3^. While most UTIs cause only mild symptoms, some ascend the urinary tract and progress to cause invasive infections such as pyelonephritis, urosepsis, and septic shock ^4, 5^. Invasive infections are associated with high morbidity and healthcare costs and can lead to septic shock and mortality ^6, 7^. Therefore, understanding the factors associated with a more severe disease outcome is critical for patient management, to decrease healthcare related costs, and improve antibiotic stewardship. Within *E. coli,* multiple distinct and deep branching phylogenetic groups (defined as phylogroups) have been identified ^8^. Of these A, B1, B2, and D are most common ^8, 9^. Phylogroups A and B1 are associated with asymptomatic carriage in the gut while phylogroups B2, D, and F predominantly cause extraintestinal infections ^10, 11^. Furthermore, there are large variations in clinical phenotypes within each phylogroup. This is linked to the presence or absence of virulence and antimicrobial resistance (AMR) genes, which can differ even between closely related strains ^9^. Consequently, UTIs can be caused by *E. coli* strains from multiple different phylogroups ^12^. Well-characterised uropathogenic *E. coli* (UPEC) virulence factors include iron uptake systems, capsular polysaccharides, immune modulators, fimbriae, and pili ^13–15^. Genes encoding these clinically important bacterial factors are associated with globally successful UPEC clones such as Sequence Types (ST) 131, ST69, ST73, and ST95^5, 13, 16^.

Among the known virulence factors, PapGII has an important role in the progression of UTI to invasive infection ^13^ and, from the perspective of invasive infection, is associated with the urinary-, and the intestinal tracts as ports of entry for *E. coli* bacteraemia ^17^. The *papGII* gene encodes one of five variants (*papGI*-*papGV*) of the adhesive tip of pyelonephritis associated pili (PAP) ^18^. Another bacterial factor which has been associated with invasive UTI is encoded by *iuc*, which is essential for the biosynthesis of the iron uptake system aerobactin ^13^.

The clinical course of a UTI and subsequent pyelonephritis and urosepsis is not only shaped by these bacterial factors, but also by human characteristics such as an effective immune response, host genetics, comorbidities, age, and gender ^2^. As such, the clinical importance of an individual bacterial factor cannot be definitively assessed without correcting for relevant patient characteristics. Despite our growing understanding of UPEC virulence-and patient risk factors, these two interconnected aspects are mostly analysed separately, and their interaction is rarely considered ^17^.

In this study, we aim to identify pathogen-and patient-specific factors associated with bacteraemia by jointly analysing genomic bacterial data and host characteristics from patients with *E. coli* positive urine-or blood culture samples. We further aim to investigate whether virulent UPEC isolates can accurately be identified in clinical routine diagnostics and to validate our findings in a second, prospectively collected cohort.

## Results

### Patient factors

To identify the most relevant patient characteristics for the progression of a UTI, we reviewed 825 clinical case charts. This included cases for which *E. coli* isolates were identified from urine (n=564) or from blood culture samples (n=261). In 106 cases, *E. coli* isolates were recovered from urine-and blood culture samples, suggesting the urinary tract as a port of entry for bacteraemia. Cases for which *E. coli* isolates were recovered from blood culture samples are henceforward referred to as ‘invasive infection’, compared to ‘non-invasive infection’ where *E. coli* isolates were recovered from urine samples, but not from blood culture samples. Patients had a median age of 75.3 years (IQR=[63.6,83.0]), a median Charlson Comorbidity Index (CCI) of 2 (IQR=[0,3]), were predominantly female (69.6%, 574/825) and 10.6% (86/812) were immunosuppressed (defined as a dose equivalent of 20 mg prednisone/day or mentioning of immunosuppression in the patient notes) (**Figure 1**, **Table 1**).

**Figure 1:**
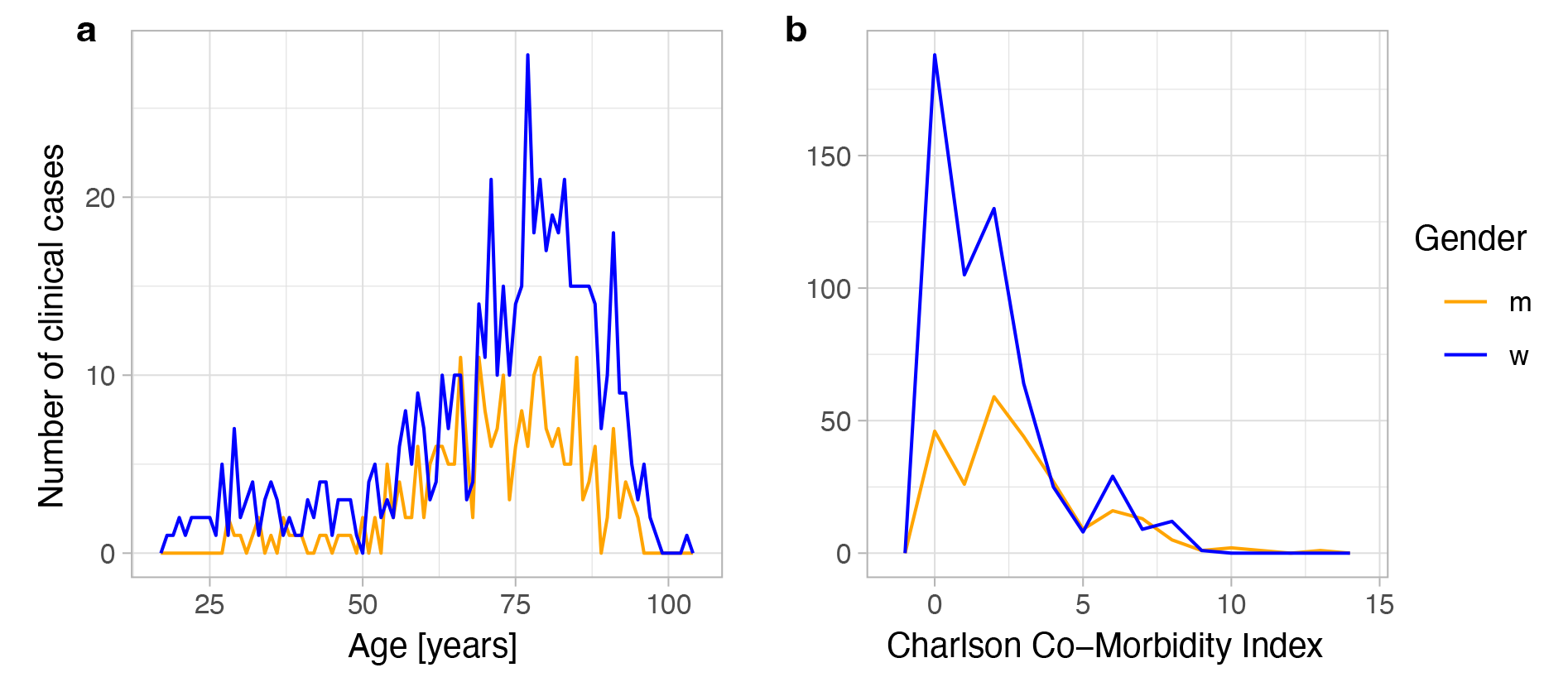
Frequency distribution of **a**: age [years] and **b**: the sum of the Charlson Comorbidity Index (CCI) for male (red) and female (turquoise) clinical cases (n=825) included in this study.

**Table 1:** Patient characteristics and outcome variables of clinical cases with 261/825 invasive or 564/825 non-invasive infection. ‘Invasive infection’ was defined as at least one positive *E. coli* blood culture, whereas ‘non-invasive infection’ was defined as at least one *E. coli* positive urine sample, with no blood culture being tested positive for *E. coli*. Some variables were unavailable for a subset of patients, resulting in varying denominators in the table.

| Variable | All cases<br>(n=825) | Cases with<br>invasive infection<br>(n=261) | Cases with non-invasive infection<br>(n=564) |
| --- | --- | --- | --- |
| Age [years] (median, [IQR]) | 75.3 [63.6,83.0] | 74.7 [63.7, 82.3] | 75.7 [63.4, 83.3] |
| Sex = male (n;%) | 251/825 (30.4) | 120/261 (46.0) | 131/564 (23.2) |
| CCI (median [IQR]) | 2 [0,3] | 2 [0,3] | 2 [0,3] |
| Immunosuppression = T (n; %) | 86/811 (10.6) | 44/256 (17.2) | 42/555 (7.6) |
| Typical UTI symptoms | 231/778 (29.7) | 77/256 (30.1) | 154/522 (29.5) |
| ICU admission | 188/821 (22.9) | 63/260 (24.2) | 125/561 (22.23) |
| 30-day all-cause mortality | 50/819 (6.1) | 21/258 (8.1) | 29/561 (5.2) |

We observed a larger fraction of rare STs (302/574, 52.6% vs. 100/251, 39.8%, p-value = 0.0010, Chi-squared test) and similar fraction of isolates belonging to carriage-associated phylogroups A, B1, and C (120/574, 20.9% vs. 42/251, 16.7% p-value = 0.196, Chi-squared test) isolated from female patients compared to male patients, whereas male patients more frequently carried strains of ST131 than female patients (49/251, 19.5% vs. 56/574, 9.8%, p-value = 0.0002, Chi-squared test) (**Figure S1**). We observed a non-significant tendency of female patients younger than 40 years (n=52) being less frequently infected with isolates from carriage associated phylogroups than female patients older than 40 years (6/52, 11.5% vs. 114/522, 21.8%, p-value = 0.118, Chi-squared test). The most frequent ST isolated from female patients younger than 40 were from ST69 (8/52, 15.4%) and ST95 (7/52, 13.5%), both of which occurred in lower frequency in female patients older than 40 (55/522, 10.5% and 22/522, 4.2%, respectively) (**Figure S2**).

The composition of phylogroups isolated from invasive and non-invasive infection was similar with 19.2% (50/261) and 19.9% (112/564) (0.200 Chi-squared test) being caused by carriage-associated phylogroups A, B1, and C. The three most common STs were the same in both invasive and non-invasive infection, namely ST69 (38/261, 14.6% and 53/564, 9.4%), ST73 (33/261, 12.6% and 51/564, 9.0%) and ST131 (34/261, 13.0% and 71/564, 12.6%). Rare STs, which overall occurred less than 20 times in our strain collection, together caused a smaller fraction of invasive infections than non-invasive infections (109/261, 41.8% vs. 293/564 52.0%, p-value = 00.0081, Chi-squared test) (**Figure S4**).

Overall cases where this information was available from the patient records, in 29.7% (231/778) of cases the patient experienced typical UTI symptoms, 22.9% (188/821) were admitted to the ICU and 6.1% (50/819) died within 30 days after the collection of the urine or blood culture sample (**Table 1**).

### Characterisation of bacterial strains

STs which comprise globally successful UPEC clones ^5, 13, 16, 19^ were frequent in our collection of isolates, namely ST131 (12.7%; 105/825), ST69 (11.0%; 91/825), ST73 (10.2%; 84/825), and ST95 (5.2%; 43/825) being the four most prevalent STs (**Figure 2A**). We further screened our isolates for the presence of *papGII* and the *iuc* operon, which have previously been identified as important factors in the progression of UTI ^13, 17^. We detected *papGII* in 20.7% of isolates (171/825) and exclusively in the ExPEC associated phylogroups B2-1 (20.6%; 29/141), B2-2 (27.0%; 97/359), D1 (32.0%; 32/100), D3 (8.0%; 2/25), and F (57.9%; 11/19). *papGII* frequently occurs within closely related strains ^13^ (**Figure 2a**). The *iuc* operon often co-occurs with *papGII* and less frequently with other *papG* variants. The complete *iuc* operon was detected in 28.8% (238/825) of all strains and in the phylogroups A (10%; 5/50) B2-1 (52.5%; 74/141), B2-2 (32.0%; 115/359), C (8.7%; 2/23), D1 (30.0%; 31/100), D3 (8%; 2/25), and F (47.4%; 9/19).

**Figure 2:**
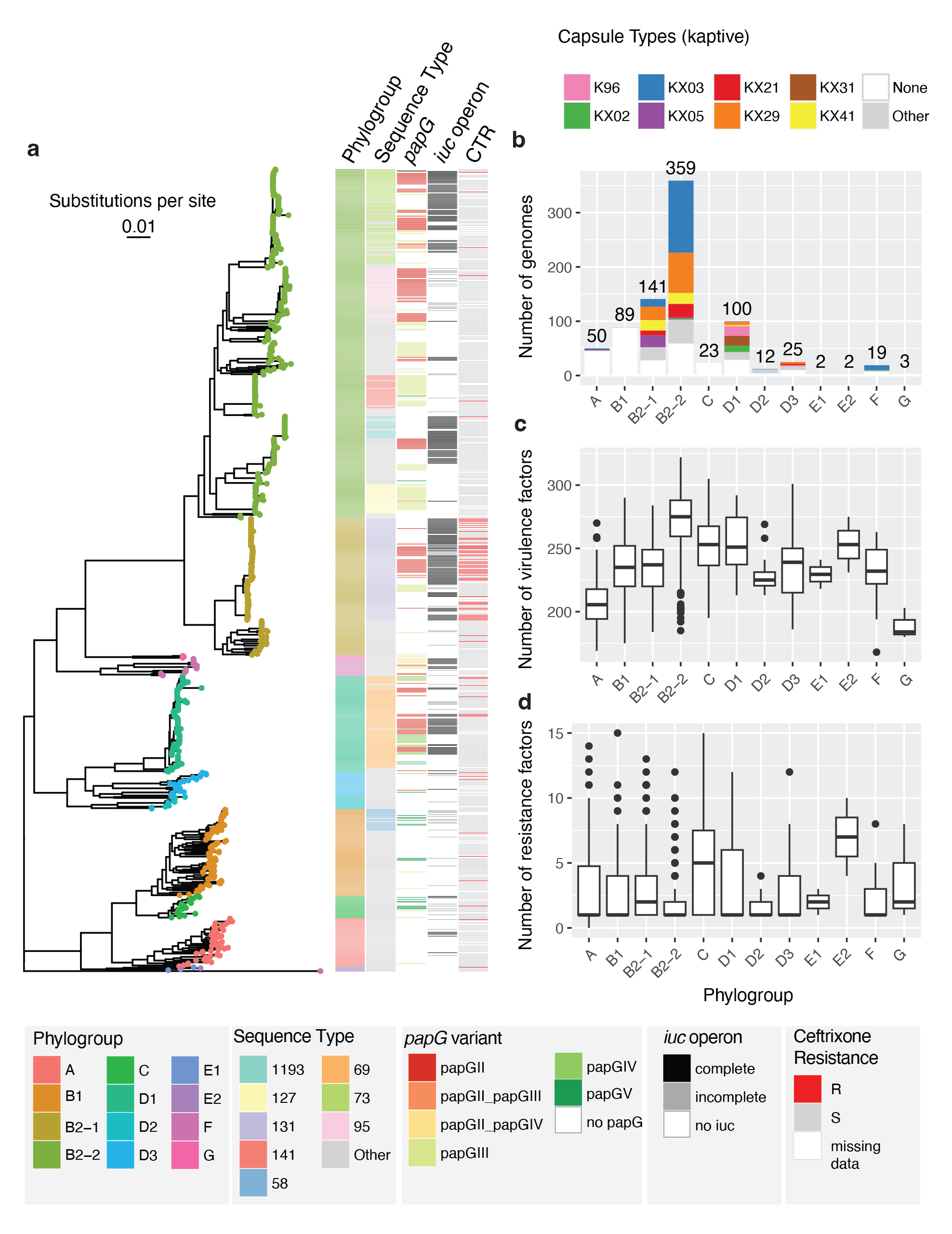
Genomic characterisation of the *E. coli* strains collected for this study (one strain per clinical case, n=825). **a:** Core genome phylogeny, phylogroup assignment, Sequence Type (ST) (8 most frequent ST are coloured, rare STs in grey), *papG* variant, occurrence of the *iuc* operon, and phenotypic ceftriaxone resistance; **b:** Frequency distribution of the *E. coli* phylogroups and their respective capsule types; **c:** Number of virulence factors detected per *E. coli* phylogroup; **d:** Number of genes associated with antimicrobial resistance per *E. coli* phylogroup.

Phenotypic ceftriaxone resistance, which is often assessed to screen for the carriage of ESBL, was rare in our dataset (9.8%; 72/737), except in isolates of ST131 (51.6%, 48/93) (**Figure 2a**).

We did not record any phenotypic resistance against meropenem (0/737), and phenotypic resistance against fosfomycin and nitrofurantoin was rare (1.5%; 8/547 and 1.1%; 6/546) and distributed throughout the phylogenetic tree (**Figure S4**). Phenotypic resistance against ciprofloxacin was more prevalent (17.0%; 125/737) and occurred most often in ST1193 (100%; 18/18) and ST131 isolates (67.7%; 63/93) (**Figure S4**).

We observed a large fraction of our isolates (79.5%; 656/825) belonging to the ExPEC associated phylogroups B2-2 (43.5%; 359/825), B2-1 (17.1 %; 141/825), D1 (12.1%; 100/825), D2 (1.5%; 12/825), D3 (3.0%; 25/825), and F (2.3%; 19/825). Only a smaller fraction of strains belonged to phylogroups associated with colonisation: A (6.1%; 50/825), B1 (10.8%; 89/825), C (2.8%; 23/825) and/or intestinal infection: E2 (0.2%; 2/825) (**Figure 2a**). We observed 63.4% (525/825) of the strains carrying genes encoding capsular polysaccharides. Strains in ExPEC-associated phylogroups most often encoded a capsule (79.1%; 519/656) whereas strains in phylogroups associated with colonisation rarely did (3.7%; 6/162). The most common capsule assignments were ‘KX03’ (30.7%; 161/525) and ‘KX29’ (20.6%; 108/525). These are the same capsule assignments, which are assigned to the well-known capsule types K1 and K5 (**Figure 2b**).

We next compared the phylogroups in terms of virulence factors and resistance genes. Of the phylogroups which included more than 10 isolates, phylogroup A strains carried the fewest virulence factors (median = 205.5, Interquartile range (IQR) = [194.25; 217.75]), while phylogroup B2-2 strains carried the most virulence factors (median = 275.0; IQR = [259.0, 288.0]) (**Figure 2c**).

Phylogroups A, B1, B2-2, D1, D2, D3, and F carried a median of one resistance gene. Of the phylogroups which cover more than 10 isolates, phylogroup C harboured the most resistance genes (median = 5; IQR=[1,7.5]), followed by B2-1 a (median = 2; IQR=[1,4]) resistance genes (**Figure 2d**).

### Genetic diversity of *E. coli* isolated from the same human host

Whereas it would be expected to find almost identical isolates in cases where *E. coli* strains were isolated from urine and blood cultures of the same patient, we found two phylogenetically distant strains with average nucleotide identity (ANI) values under 99.9% in 12.3% (13/106) of cases (**Figure S5a**). To compare the strain diversity within urine and blood culture samples, we picked ten colonies for single colony sequencing each from urine and blood culture samples of three patients with *E. coli* bacteraemia. In one of the three urine samples, we observed phylogenetically distant strains, sharing under 99.9% ANI and displaying 71,193 - 80,916 single nucleotide variants (SNV) in pairwise comparisons. When pairwise examining closely related strains from the same sample (over 99.9 ANI), we identified a higher number of SNVs (median=3, IQR=[2,5]) for strains co-isolated from urine samples than for strains co-isolated from blood culture samples (median=2, IQR=[0,2], p-value = 0.00033, Mann Whitney U test (**Figure S5b**).

### Bacterial genome wide association study

To identify bacterial factors promoting UTI progression, we performed a bacterial genome wide association study (GWAS), including one isolate per clinical case and using ‘bacteraemia’ as clinical endpoint (=’invasive infection’). If multiple isolates per clinical case had been collected, we chose isolates recovered from blood culture samples over isolates recovered from urine samples, as these caused the invasive infection. ‘Bacteraemia’ was defined as at least one positive *E. coli* blood culture, compared to ‘non-invasive UTI’ which was defined as at least one *E. coli* positive urine sample, with no blood culture being tested positive for *E. coli* within seven days.

We identified 13 genes to be significantly associated with invasive infection (**Figure 3a**, **Supplementary Table S1**). The most significant hit corresponds to *papG* (89 unitigs (i.e. kmers of variable length)). 70/89 of these *papG* unitigs had the highest ANI to the *papG* variant *papGII* and 15/89 additional significant unitigs were equally similar to *papGII* and *papGIII*. 2/89 unitigs had the highest similarity to *papGIII*, whereas 1 unitig each matched most closely to *papGI* and *papGIV* (**Figure S6**).

**Figure 3:**
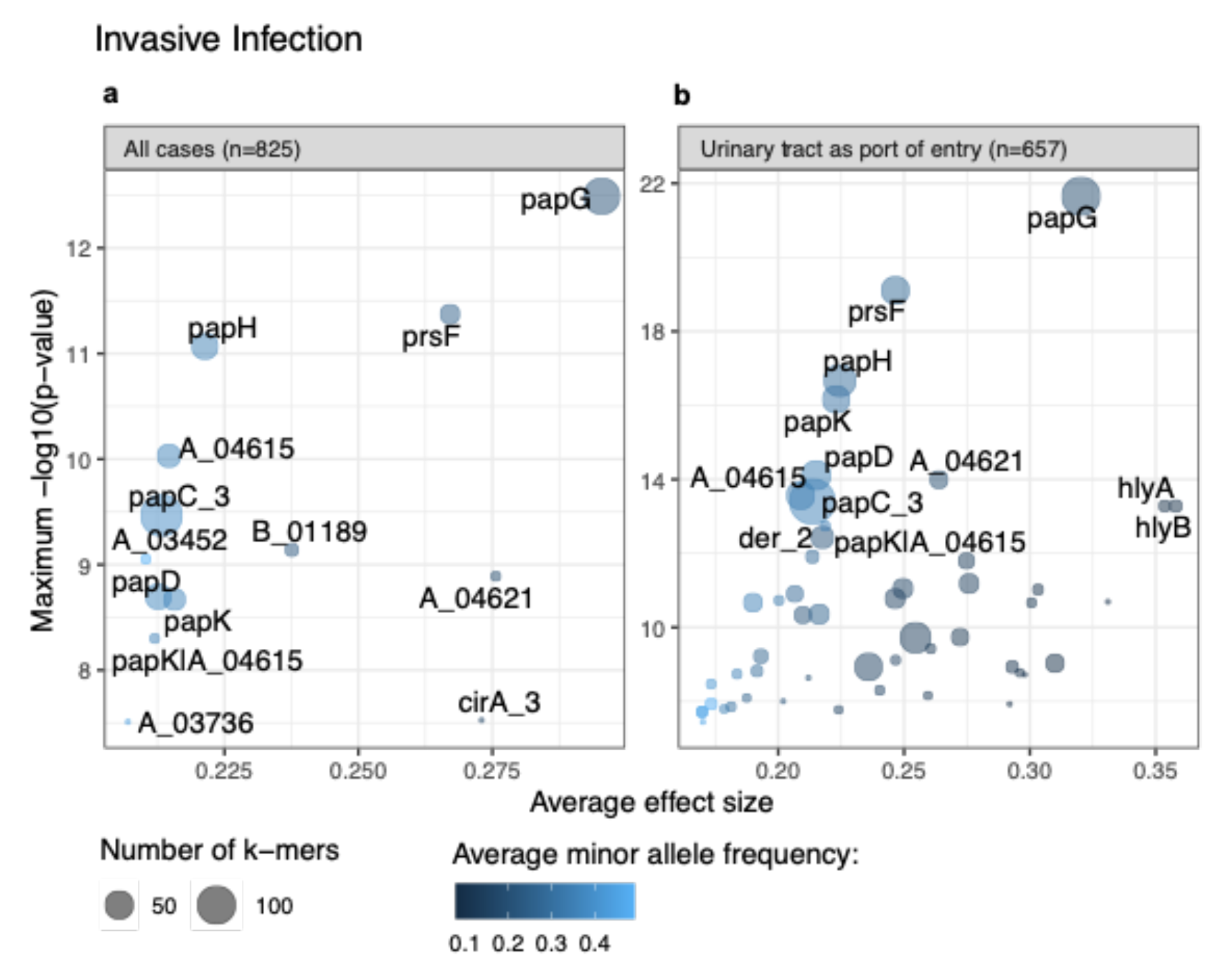
Significance level and average effect size of genes with mapping unitigs identified as significant in a bGWAS **a:** including all clinical cases (n=825) and **b:** including cases for which the port of entry for bacteraemia could be assigned to the urinary tract. In B only genes with a maximum - log10(p-value) > 11 are labelled. Genes labelled with ‘A_’ and ‘B_’ correspond to annotated genes of the strains ‘100033-19’ and ‘100888-20’ in our collection, respectively.

We identified nine additional genes of the *pap* operon to be associated with invasive infection: *prsF* (*papF*), *papH*, *papC*, *papK*, a gene annotated as *papJ|papK* and *100888-20_01189* and *100033-19_04615* (both identified as *papJ* using blastp) and *100033-19_04621* (identified as *papI* using blastp). Furthermore, our GWAS identified unitigs mapping to the gene *cirA* as associated with invasive infection. CirA is embedded in the outer membrane, is postulated to participate in iron transport, and serves as a receptor for the colicins IA and IB ^20^. For two genes (annotated as *100033-19_03452* and *100033-19_03736*), which also showed significant association with invasive infection, comparison to the EggNOG database identified NlpC/P60 family domains. Using NCBI blastn, both genes showed 100% sequence similarly with 100% query coverage to genes encoding C40 family peptidases and phage tail proteins (**Supplementary Table S1**).

Of the clinical cases which carried a *papGII* positive *E. coli* strain, 56.7% (97/171) developed invasive infection, whereas 25.1% (164/654) of *papGII* negative clinical cases developed bacteraemia. Using *papGII* as a predictor for invasive infection had a sensitivity of 37.2%, a specificity of 86.9%, a positive predictive value of 56.7% and a negative predictive value of 74.9%.

In a further analysis, we focused on UTI as the port of entry for sepsis. In a second bGWAS we therefore excluded all invasive cases for which *E. coli* genomes were not available from isolates recovered from a blood culture and a urine sample, or where the *E. coli* isolate from the urine sample was phylogenetically distant to the blood culture isolate. In line with the previous GWAS, the genes of the *pap*-operon are most significantly associated with invasive infection (**Supplementary Table S2**, **Figure 3b**). In addition to the genes identified in our previous GWAS, we observed 41 more genes to be significantly associated with invasive infection. These include genes involved in basic cell metabolism, genes of unknown function, remnants of phages and other mobile genetic elements as well as known *E. coli* virulence factors. These include *hlyA* (encoding an alpha-hemolysin ^21^), *hlyB*, *hlyD* (involved in the hemolysin transport ^22, 23^), and *cbtA* and *cbeA* (encoding the toxin and antitoxin part of a type IV toxin-antitoxin system, respectively ^24, 25^).

In this smaller, more focused cohort, 41.3% (52/126) of cases which were infected with a *papGII* positive *E. coli* strain developed invasive infection, compared to 7.7% (41/531) of *papGII* negative clinical cases. Using *papGII* as a single predictor for invasive infection had a sensitivity of 55.9%, a specificity of 86.9%, a positive predictive value of 41.3% and a negative predictive value of 92.3%.

### *papGII* is associated with invasive infection when correcting for patient characteristics

To concurrently assess the impact of pathogen and patient-specific factors on the progression of UTI, we used a generalised linear model with ‘bacteraemia’ as primary endpoint and ‘typical UTI symptoms’, ‘admission to ICU’ and ‘30 day all-cause mortality’ as secondary endpoints. As bacterial factors we included *papGII* carriage, being the most prominent hit identified in our GWASs. With the aim of identifying bacterial virulence and to correct for differences in AMR, we have additionally included phenotypically tested resistance against ceftriaxone as a bacterial factor in our model.

Across all cases, *E. coli* strains encoding *papGII* were significantly more likely to be involved in invasive infection (OR 5.27, 95% CI = [3.48,7.97], p-value < 0.001) (**Figure 4**). We observed clinical cases with immunosuppression (OR 2.82, 95% CI = [1.64,4.87], p-value < 0.001) and male patients (OR 3.48, 95% CI = [2.39,5.06], p-value < 0.001) being more likely to be associated with invasive infection (**Figure 4**).

**Figure 4:**
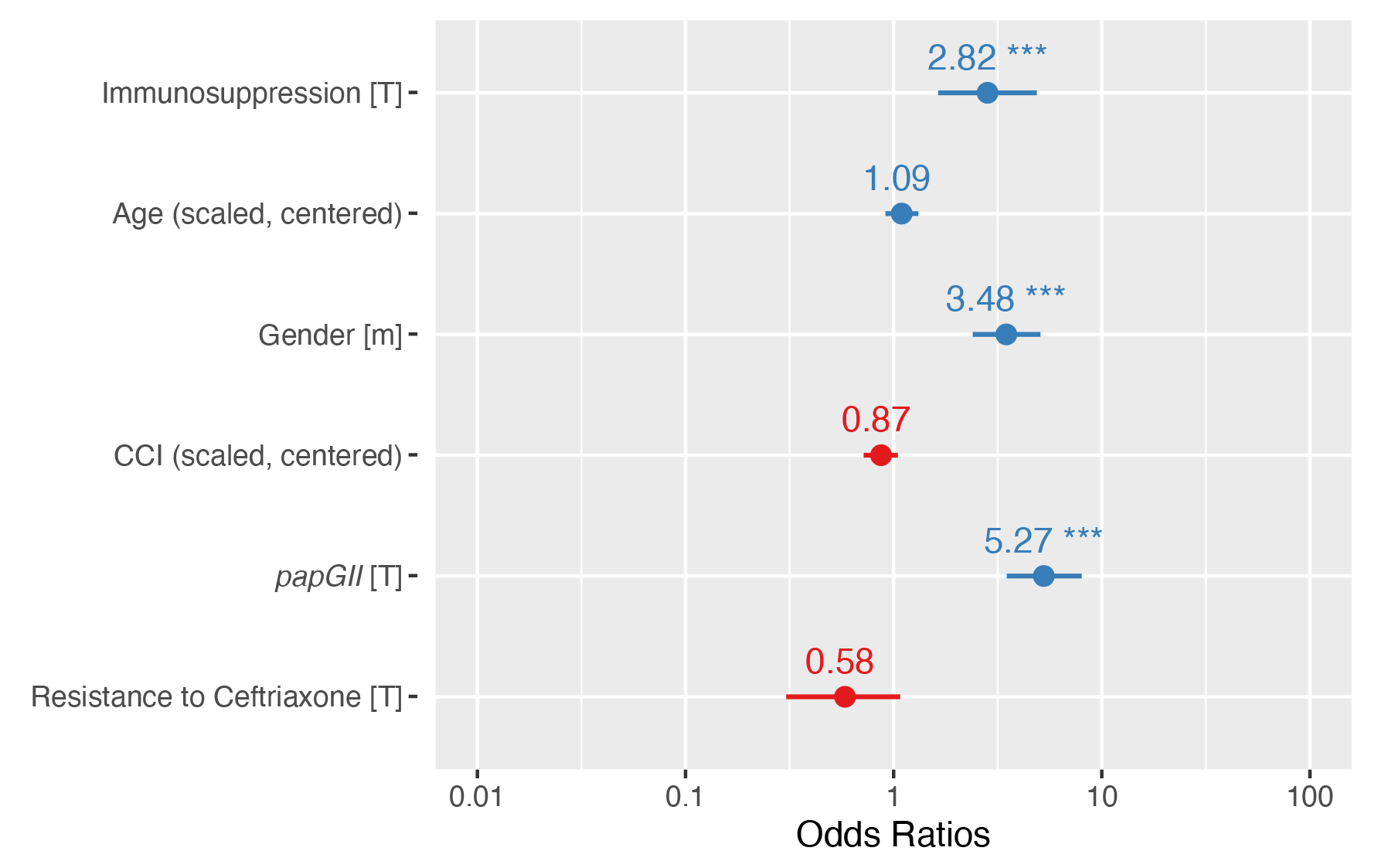
Odds ratio estimates with 95% confidence intervals for invasive infection using the generalised linear model (GLM). *n =* 751 complete observations with 210 events. CCI = Charlson Comorbidity Index

Although not significant, and to a lesser extent, we also observe a tendency of *papGII* encoding *E. coli* strains to be more likely involved in (i) clinical cases with typical UTI symptoms (OR 1.44, 95% CI = [0.97,2.13], p-value = 0.072) (**Figure S7a**) and (ii) clinical cases involving an ICU stay (OR 1.45, 95% CI = [0.96,2.20], p-value = 0.078) (**Figure S7b**) than *E. coli* strains which do not encode *papGII*. Clinical cases with *papGII* positive strains showed a significantly higher concentration of C-reactive Protein (CRP) (median=115.6mg/l, IQR=[47.8,206.8] vs. median=41.0mg/l, IQR=[11.2,113.5], p-value<0.001, Mann Whitney U test)(**Figure S8a**), a higher leukocyte count in blood samples (median=10.9×10^9^/l, IQR=[7.7,14.4] vs. median=9.1×10^9^/l, IQR=[6.5,12.2], p-value<0.001, Mann Whitney U test)(**Figure S8b**) and a significantly higher ratio of leukocyte/bacterial cell counts in urine samples, compared to cases with a *papGII* negative isolate (median=0.19, IQR=[0.06,0.68] vs. median=0.10, IQR=[0.02,0.45], p-value = 0.0018, Mann Whitney U test) (**Figure S9a**). Urine samples of cases for which a *papGII* positive strain was isolated, showed a tendency towards a higher bacterial cell count (median = 7194/uL, IQR = [1473,12914] vs. median = 5224/uL, IQR=[764,11218], p-value = 0.044, Mann Whitney U test) (**Figure S9b**), were less frequently tested positive for nitrite than urine samples of cases with *papGII* isolates (**Figure S9c**), although these differences were only marginally or not significant (34.4% 54/157 vs. 42.8%; 246/575, p-value = 0.07, Chi-squared test). No difference in the ratio of erythrocytes/bacterial cell counts was detected between urine samples with *papGII* positive and *papGII* negative *E. coli* strains (p-value = 0.28, Mann Whitney U test) (**Figure S9d**).

We found no evidence of a *papGII* specific association with 30-day mortality (p-value = 0.31) (**Figure S7c**). Patients 40 years and older were more often infected with *papGII* negative strains (610/760, 80.3%) compared to patients younger than 40 years (44/65, 67.7%) (p-value = 0.03, Chi-squared test), whereas there was no evidence for varying *papGII* frequencies between male and female patients (p-value = 0.40, Chi-squared test), or between immunosuppressed and immunocompetent patients (p-value = 0.37, Chi-squared test) (**Figure S10**).

Older patients were less likely to experience typical UTI symptoms (OR 0.82, 95% CI = [0.70,0.97], p-value = 0.018) (**Figure S7a**) and more likely to die within 30 days of diagnosis (OR 2.54, 95% CI = [1.51,4.28], p-value < 0.001). Patients with an increased CCI (OR 1.68, 95% CI = [1.28,2.16], p-value < 0.001) were more likely to die within 30 days after the *E. coli* diagnosis (**Figure S7c**).

### No evidence for *papGII* specific peak in MALDI-TOF mass spectra

Our results and previous studies highlight the importance of *papGII* in invasive *E. coli* infection. Its early detection in clinical diagnostics would be desirable and give evidence towards a severe progression of an ongoing UTI. MALDI-TOF MS is the most widely used tool for bacterial species identification in routine diagnostics and we therefore aimed to assess whether there are *papGII* specific signals in MALDI-TOF MS spectra. We did not identify any *papGII* specific signal, nor a peak being associated with the absence of any *papG* variant (**Figure S11**).

As *papGII* presence is associated with the ExPEC phylogroups B2-1, B2-2, D1, D2, D3, and F, we further investigated whether the *E. coli* phylogroups can be distinguished by MALDI-TOF MS. We identified a mass shift from around 9711 Da to 9739 Da, which uniquely distinguishes the phylogroups B2-1, B2-2, and F from all other phylogroups (**Figure S12**). This mass shift has previously been identified as two different mass alleles of the acid stress chaperone HdeA ^26^. From MALDI-TOF mass spectra (**Figure S12**) and predicted from genomic data (**Figure S13**) we observe the HdeA mass allele at 9712 Da in the ExPEC associated phylogroups B2-1, B2-2, and F and the mass allele at 9740 Da in the phylogroups A and B1 which are associated with a colonisation clinical phenotype. However, the ExPEC associated phylogroups D1, which also carries *papGII*, and D3 also encoded HdeA with a mass of 9740 Da. Strains of the phylogroup D2 were observed to encode HdeA of either 9712 Da or 9740 Da (**Figure S12** and **Figure S13**).

### *papGII* can be identified by qPCR directly from urine samples

As an alternative diagnostic method, we designed and validated qPCR primers and probes to detect *E. coli* strains encoding *papGII*. After verifying the functionality of our primers and probes (**Figure S14-S18**, **Supplementary PCR Data**), we prospectively screened 1,657 urine samples at two health care centres (n=529 at Centre 1 and n=1128 at Centre 2).

Of the 261 samples that tested culture positive for *E. coli* in Centre 1 in the routine diagnostic, a *gapC* signal was detected in 98.8% (258/261) using our PCR assay. A *gapC* signal was further recorded in 36.2% (97/268) of samples, for which no *E. coli* was reported in clinical routine diagnostics, possibly reflecting either the increased sensitivity of our qPCR assay compared to culture-based detection of *E. coli*, or unspecific amplification. *papC* was detected in 267/529 samples, 63.7% (170/267) of which were culture positive for *E. coli*. Finally, *papGII* was detected in 40/529 urine samples which were all *E. coli* culture positive. We further evaluated which clinical cases for which *E. coli* had been cultured in routine diagnostics (n=244, excluding multiple samples from the same patient) developed an invasive infection. An *E. coli* positive blood culture was recorded in 8.3% (3/36) of cases for which a *papGII* encoding *E. coli* had been identified in the urine sample, compared to 4.3% (9/208) of cases for which an *E. coli* with no *papGII* had been identified in the urine sample (p-value 0.54, Chi-squared test).

In comparison to Centre 1 where all urine samples were prospectively screened, we exclusively screened culture positive *E. coli* isolates at Centre 2 (n=1,106). When including one sample per patient only (n=871), we detected *papC* in 33.8% of cases (293/868, 3 measurements were excluded as the positive control (rpoD) did not test positive) and *papGII* in 16.4% of cases (143/871). Patients with a *papGII* positive strain developed more frequently an invasive infection than patients with a *papGII* negative strain (11.9% (17/143) vs. 6.5% (47/728), p-value = 0.036, Chi-squared test).

## Discussion

In this study we concurrently analysed bacterial genomic factors and patient characteristics and highlighted the importance of *papGII* in invasive UTI infections. Consistent with previous studies, we identified *papGII* in globally successful UPEC lineages ^13^.

*papGII* often co-occurs with *iuc* and rarely occurred in strains which were resistant to ceftriaxone except within the globally disseminating ESBL UPEC lineage ST131. We observed over 60% of our strains and over 95% of our ExPEC phylogroup strains carrying genes which encode for polysaccharide capsules. This is a substantially larger fraction than has previously been reported in *E. coli* RefSeq sequences where less than 25% of assemblies carried the *kpc* locus ^27^. Capsular polysaccharides facilitate Gram-negative bacteria to evade the innate host immune response ^28^. For example, the *E. coli* capsules K1 and K5 have been shown to do so by molecular mimicry because identical polysaccharides are present on human cells ^29^. The increased proportion of polysaccharide capsules in our collection gives further evidence on the importance of capsules for *E. coli* to infect the human urinary tract. However, neither the data collected in this study nor those from previous studies assessing *E. coli* bloodstream infection ^13, 17^ suggested an association of these capsule loci with severe progression of UTI to invasive infection.

In a subset of samples, we identified the *E. coli* strain isolated from the urine sample being unrelated to the *E. coli* strain isolated from the bloodstream from the same clinical case. Possible explanations for this observation are (i) a multi-strain infection of the urinary tract or (ii) a different port of entry to the bloodstream than the urinary tract. In a continuous analysis and in line with previous studies ^30, 31^, we identified strains of different phylogroups isolated from the same urine sample which supports explanation (i). However, as we examined multiple colony picks from the same urine sample only for a small number of cases, further studies are required to assess the within-host *E. coli* strain diversity infecting the urinary tract and their dynamics in disease progression.

In line with a previous study ^13^ we identified genes of the *pap* operon, and most significantly *papGII*, as being associated with invasive infection in both our bacterial GWASs. The gene *papGII* encodes one of multiple variants (PapGI - V) of the adhesin tips of Pap pili, and binds to the globoseries of glycosphingolipids, more specifically Gb5 (GalNAcα1-3-GalNAc3Galα1- 4Galβ1-4GlcCer) ^17, 32^ of human uroepithelial and kidney cells ^33, 34^ and can modulate the host immune response ^35^. A previous study suggests that virulent UPEC lineages emerged after the independent horizontal acquisition of pathogenicity islands encoding *papGII* ^13^. Factors associated with horizontal gene transfer (HGT) (e.g. phage/prophage associated proteins) and *E. coli* toxins, were identified as associated with invasive infection in our bGWASs, although less significantly than *papGII*. These genes potentially originate from the horizontal acquisition of the pathogenicity island encoding the *papGII*, which carries varying gene content, occasionally including these toxins (*hlyA*, *hlyB*, *hlyD* on PAI Type II, III and VI)^13^.

The crucial role of *papGII* in invasive infections is further supported by our GLM, where we concurrently correct our analysis for important patient characteristics as well as resistance against ceftriaxone. Correcting for patient characteristics is a crucial step to estimate bacterial virulence. Without such analyses it is not possible to assess whether an observed increase in virulence is caused by a bacterial genetic factor or might arise from patient confounding factors. Our analysis also identified the higher likelihood of male patients developing invasive infection, which can presumably be explained by the increased occurrence of uncomplicated UTI in female patients ^36^. In our GLM we did not consider other ports of entry for sepsis than the urinary tract, which certainly is a limitation of the current study as this might influence the clinical outcomes. The urinary tract has previously been determined as the entry point in 50- 60% of *E. coli* sepsis ^17^. As *papGII* binds to human uroepithelial and kidney cells ^37, 38^, its impact on patient outcome might be even larger when exclusively examining patients with urosepsis, compared to all bloodstream infections.

In this study we classified clinical cases into ’invasive’ (positive blood culture) and ‘non-invasive’ (positive urine culture, no positive blood culture) infections based on microbiological endpoints. This classification is not always consistent with a clinical assessment. A more refined classification of cases as well as considering antimicrobial treatments and co-morbidities in more detail would further improve the risk assessment of bacterial and patient factors.

We examined whether there are MALDI-TOF MS peaks which are specific for *E. coli* strains carrying *papGII* and which could be used to detect these virulent UPEC clones in clinical routine diagnostics. The *papGII* protein weighs 37,667 Daltons ^39^ and lies beyond the mass range of MALDI-TOF MS devices routinely used for bacterial species identification. Unfortunately, no alternative peak was observed which could serve as a surrogate marker. We did, however, observe a previously described mass shift ^40^ of the acid stress chaperone *HdeA* which is specific for the ExPEC phylogroups B2-1, B2-2 and F but fails to discriminate the ExPEC phylogroup D1-3 from the carriage associated phylogroups A, B1 and C. Although in this simple analysis no single signal was observed which unambiguously identifies *papGII* positive strains from MALDI-TOF mass spectra, more elaborate statistical analysis might still reveal a combination of peaks or intensity patterns allowing for the identification of such strains.

As an alternative approach to screen patient samples for *E. coli*, the *pap* operon and *papGII*, we designed qPCR primers and probes for three targets and tested their performance when applied directly to urine samples. The advantage of this method is the short turnaround time, which allows results to be obtained a few hours after sampling as the cultivation step can be omitted. For the primer pairs for the *E. coli* marker *gapC* and the pap-operon marker *papC*, amplifications were recorded in few samples for which no positive *E. coli* culture was reported. These could correspond to non-specific amplifications or, as we hypothesise, indicate an increased sensitivity of qPCR compared to culture-based detection. To be applicable in routine diagnostics, this method would have to be further validated, including samples with other *Enterobacteriaceae*.

In both centres where we prospectively screened *E. coli* strains infecting the urinary tract, we observed the relative frequency of invasive infection to be >1.8 fold higher for *papGII* positive strains compared *papGII* negative strains which is in line with our first cohort included in our bGWAS. These findings from an independently collected patient cohort further substantiates the importance of *papGII* in invasive infections, which we have previously highlighted by analysing bacterial genomic data and important patient characteristics. This study exemplifies the importance of including patient data to assess the potential virulence of a bacterial pathogen.

## Methods

### Ethics

The collection and analysis of strains and patient data was approved by the ‘Ethikkommission Nordwest-und Zentralschweiz’ (EKNZ) (BASEC-Nr. 2019-00748).

### Patient data collection

To assess which patient characteristics impact the progression of a UTI, we reviewed 825 clinical case charts. Demographic and clinical data was systematically collected for patients with an *E. coli* isolated from blood or urine between 04/2018 and 02/2020 at the University Hospital Basel by retrospective electronic chart review. The University Hospital Basel is a tertiary healthcare centre with more than 750 beds in a low endemic region for extended spectrum β-lactamase (ESBL)-producing bacteria (22). Inclusion criteria were: patients for which at least one isolated bacterial colony was collected from urine or blood culture samples, identified as *E. coli* by MALDI-TOF MS (Bruker Daltonics, Bremen, Germany) or using biochemical assays on the Vitek2 (bioMérieux, Marcy-l’Étoile, France); no negative statement for the hospital’s general research consent as approved by the ethical committee. The following clinical data was recorded: presence of typical urinary tract symptoms (dysuria, increased urinary frequency, urgency ^41^), mention of *E. coli* infection as a medical diagnosis, and laboratory results (including leukocytes, C-reactive protein, and urine flow cytometry) on the day (+/-1) of urine and/or blood sample collection, as well as the requirement for antibiotic treatment. Information on the need for and duration of hospitalisation, admission to the ICU and in-hospital mortality were extracted. Additionally, we determined 30-day all-cause mortality, defined as a death occurring for any reason within 30 days of the collection of the urine or blood culture sample. Relevant comorbidities were recorded according to the Charlson Comorbidity Index ^42^. Immunosuppression defined as corticosteroid dose equivalent to prednisone 20 mg daily, active haematological malignancy history of hematopoietic stem cell transplant or solid organ transplant or absolute neutrophil count <500 cells/µl. We summarised patient demographic and clinical data into units of clinical cases, which were defined as a unique hospital stay and included data collected from the same patient between the hospital entry and the hospital exit date.

### Definition of ‘Invasiveness’

*E. coli* strains were defined as ‘invasive’ if isolated from a blood culture or if a blood culture from the same clinical case was diagnosed positive for *E. coli* within seven days before or after the collection of the *E. coli* positive urine sample. Similarly, the term ‘invasive infection’ was used to describe clinical cases with at least one *E. coli* positive blood culture sample, whereas ‘non-invasive’ cases described all other cases in our cohort.

### Bacterial isolate collection and whole genome sequencing

*E. coli* isolates (n=1,076) were prospectively collected at the University Hospital Basel from 825 clinical cases from urine (n=789), blood culture (n=286), and deep tissue samples (n=1) from 04/2018 to 02/2020, aiming for a balanced dataset between invasive and non-invasive infections. Urine was cultured on 5% Columbia sheep blood (COS) agar plates (Becton Dickinson, New Jersey, USA) and Chrom ID plate (Becton Dickinson, New Jersey, USA) for a maximum of 48 hours. Blood cultures were incubated in aerobic and anaerobic flasks for a maximum of six days using the Virtuo system (bioMérieux, Marcy-l’Étoile, France). Bacterial isolates were identified as *E. coli* in routine diagnostics using the microflex Biotyper MALDI-TOF MS system (Bruker Daltonics, Bremen, Germany). For whole genome sequencing, isolates were grown on COS agar (bioMérieux, Marcy-l’Étoile, France) and DNA was extracted using the QIACube with the QIAamp DNA Mini Kit (QIAGEN, Hilden, Germany). After quality control of the DNA by Tapestation (Agilent, Santa Clara, USA), tagmentation libraries were generated as described by the manufacturer (Illumina DNA Prep Kit, Illumina, San Diego, USA). The genomes were sequenced using a 2 x 300 base pairs V3 reaction kit on an Illumina MiSeq or using a 2 x 150 base pairs on an Illumina NextSeq500 instrument. Raw reads of all isolates are publicly available via the European Nucleotide Archive (Project Accession PRJEB55855).

### Phenotypic antimicrobial susceptibility testing

Antimicrobial susceptibility testing (AST) was performed in routine diagnostics using the Vitek2 system (*Enterobacteriaceae* AST Card, bioMérieux, Marcy-l’Étoile, France) or using gradient diffusion strips (Liofilchem, Roseto degli Abruzzi, Italy) and the measurements were interpreted according to EUCAST clinical breakpoints (v.9.0) into the categories ‘Susceptible’, ‘Intermediate’ or ‘Resistant’.

### Comparative genomic analysis

We trimmed the raw reads using Trimmomatic (v0.38) ^43^, generated assemblies using Spades^44^ via unicycler (v0.3.0b) ^45^, and polished them using pilon (v1.23) ^46^. We examined the following features to ensure the quality of the sequence data and assembly: average read quality (median = 98.4; range = [56.4, 99.5]) and depth (mean = 74.6X; range = [12.9X, 328.8X]), %G+C content (median = 50.6; range=[50.3-51.1]), genome size (median = 5.0 megabases (MB); range=[4.4-5.9]). The purity of the sample was assessed using MetaPhlan^47^ and genomes were annotated using prokka (v1.13) ^48^. Bacterial species identification was confirmed using ribosomal Multi Locus Sequence Typing (rMLST) ^49^, where three strains were identified as *Escherichia marmotae* and one as *Escherichia ruysiae*. These were excluded from further analysis. We screened all assemblies for the occurrence of previously described UPEC virulence (EcVGDB, ^13^) and resistance factors (NCBI, ^50^) using abricate (v0.8.10) (https://github.com/tseemann/abricate) and >95% coverage and >95% identity thresholds. On the basis of genomic Mash distances the *E. coli* phylogroups have recently been suggested to be split up, resulting in 14 phylogroups, including two which correspond to strains of the genus *Shigella* ^11^. In order to assign our genomes to these phylogroups, we calculated the mash (v2.2) ^51^ distance to the medoid reference genomes of each phylogroup ^52^, except for phylogroup C, where an alternative reference genome was used (GCF_001515725.1), as the published medoid reference genome clustered within phylogroup B1. We chose an alternative reference genome for phylogroup C from the Microreact project (https://microreact.org/project/10667ecoli/c38356ec) belonging to phylogroup C according to ‘PCR Phylogroup’ and ‘Mash-Screen-Phylogroup’ and having the highest ’Total Score’ and ’Sequence Score’. We assigned the phylogroup of the closest reference genome to each queried genome using a cut-off of 0.04 mash distance ^11^. We identified the *E. coli kpc* loci and capsule types using fastKaptive (v0.2.2) ^27^, considering the best hit per genome if a minimal coverage of 80% to a reference loci was detected and no reference gene was missing. To link the fastKaptive assignment to phenotypically assigned Capsule Types, we ran reference genomes for the well-known Capsule Types K1 (CP003034.1) and K5 (CP022686.1) through fastKaptive, which were assigned as ‘KX03’ and ‘KX29’, respectively. We determined the O-and H antigens as well as the Multi Locus Sequence Type (MLST) via srst2 (v0.2.0) ^53, 54^. We assessed the Average Nucleotide Identity (ANI) between sequenced isolates using fastANI (v1.32) ^55^. We used Panaroo (‘sensitive’ mode) (v1.2.7) ^56^ to identify the core genome which we aligned using mafft (v7.467) ^57^ and used RaxML (`GTRCAT` approximation) (v8.2.8) ^58^ to construct a phylogenetic tree from this alignment. The output from panaroo includes a nucleotide alignment for each gene. The alignment of the gene *hdeA* was translated to amino acid sequences using transseq ^59^. The signal peptide ^60^ was removed, before predicting the pass of the HdeA protein using protparam ^61^. To investigate the diversity of *E. coli* isolates within a sample we sequenced ten colonies picked from three blood culture samples and three urine samples each. For pairwise comparison of isolates recovered from the same patient sample, we used the variant caller Freebayes (v1.2.0) ^62^ via snippy (minimum mapping quality = 60, minimum basequality = 30, minimum coverage = 30, minimum proportion for variant evidence = 0.95,v4.3.6) (https://github.com/tseemann/snippy).

### Bacterial Genome Wide Association Study

To identify bacterial factors associated with invasive infection, we used pyseer (v1.3.9) ^63^. We used unitigs as inputs and invasive infection (i.e. bacteraemia) as clinical endpoint. Unitigs represent non-redundant sequence elements of variable lengths, which we had previously constructed via unitig-counter (v1.0.5) ^64^. A minor allele frequency threshold of 5% was used. One isolate per clinical case (n=825) was included in the analysis. We treated each clinical case as a single event and included one *E. coli* genome per case in the analysis. If multiple strains per clinical case had been isolated, we chose isolates recovered from blood culture samples over those recovered from urine samples, as these caused the invasive infection. If there were multiple strains isolated from the same material, we included the isolates recovered at the earliest time point. We used random effects to correct for population structure (‘-lmm’ mode) by providing a similarity-matrix acquired from the core genome phylogeny (constructed as described above). Unitigs were mapped against the genome annotations of all strains (n=825). In order to identify the *papG* variant against which the unitigs mapped, we compared these to the reference sequences for *papGI*, *papGII*, *papGIII*, *papGIV,* and *papGV* ^13^ using fastANI (v1.32) ^55^.

To focus on the urinary tract as a port of entry for invasive infection, the analysis was repeated excluding invasive strains for which no matching urine isolate was identified in our dataset (168 strains excluded, 657 strains remaining including 93 causing invasive infection and 564 not causing invasive infection).

### Generalised Linear Models

We evaluated the impact of *papGII* on clinical outcomes by building generalised linear models, correcting for important patient characteristics. Such analyses are important to assess the potential of *papGII* as a diagnostic marker for virulent infection. The clinical outcomes analysed in this study included (i) invasive infection (defined as at least one *E. coli* positive blood culture sample, see above) (ii) experience of typical UTI symptoms, (iii) admission to an intensive care unit (ICU), and (iv) all-cause mortality within 30 days of *E. coli* diagnosis. Clinical outcomes were examined for an association with *papGII* carriage and phenotypic resistance to ceftriaxone of the infecting *E. coli* strain, age, gender, immunosuppression (defined as a dose equivalent of 20mg prednisone/day or mentioning of immunosuppression in the patient notes), and Charlson Comorbidity Index (CCI) ^42^. We corrected for resistance to ceftriaxone, as we aimed to investigate bacterial virulence and not AMR impacting patient outcome. If multiple strains per clinical case had been isolated, we chose *papGII* carriage and ceftriaxone resistance of strains isolated from blood culture samples, over strains isolated from urine samples. If multiple strains had been isolated from the same material, we chose the *papGII* carriage and ceftriaxone resistance of the strain isolated at the earliest time point. In order to compare the effect size between the included variables, we scaled and centred the numerical variables ‘age’ and ‘CCI’. All outcomes were binary and were analysed using generalised linear models (GLM) with binomial error distribution. Statistical analyses were performed in R (v 3.7).

### MALDI-TOF MS

We aimed to identify virulent *E. coli* strains using MALDI-TOF MS. We acquired MALDI-TOF mass spectra for a subset of *E. coli* isolates (n=303). This subset represents strains encoding *papGII* (n=78) and strains which did not encode *papGII* (n=225), as well as representative isolates of the phylogroups A (n=19), B1 (n=26), B2-1 (n=48), B2-2 (n=135), C (n=7), D1 (n=42), D2 (n=5), D3 (n=13), and F (n=8). Each strain was measured in quadruplicate on two MALDI-TOF MS devices including a Microflex Biotyper ‘smart’ (Bruker Daltonics, Bremen, Germany) and an Axima Confidence (Shimadzu, Ngoyo, Japan) using direct smear method and overlaying with 1μl formic acid (25%) and 1μl cyano-4-hydroxycinnamic acid (CHCA) matrix solution.

Mass spectra acquired on the Axima Confidence were exported as ‘mzXml’ and mass spectra acquired on the microflex Biotyper as ‘fid’ files and both were further processed in R using the packages MALDIQuant and MALDIQuantForeign ^65^: Mass spectra were trimmed to a mass range of 4000 - 20,000, the intensity was transformed (‘sqrt’) and smoothed (method="SavitzkyGolay",halfWindowSize=20), the baseline was removed (method="SNIP", 40 and 160 iterations for spectra acquired on the microflex Biotyper or the Axima Confidence, respectively) and the intensity was calibrated (method=”median”) before peaks were detected ("SuperSmoother", halfWindowSize= 20, SNR=2). Peaks were calibrated by aligning the mass spectra to 23 conserved masses (4364.4 Da, 5095.8 Da, 6371.5 Da, 6446.3 Da, 6541.7 Da, 7273.4 Da, 7288.8 Da, 8499.9 Da, 9006.4 Da, 9704.3 Da, 10430.2 Da, 11564.2 Da, 11580.4 Da, 11735.4 Da, 12769.5 Da, 13133.1 Da, 13540.9 Da, 14126.4 Da, 14875.2 Da, 15281.0 Da, 15768.9 Da, 17603.2 Da, 17711.4 Da) and 1000 ppm tolerance in both directions. All scripts can be accessed via GitHub (https://github.com/acuenod111/UPEC). All raw mass spectra and processed peak lists can be accessed via the Open Science Foundation (https://osf.io/vmqc5/).

### Real Time Polymerase Chain Reaction (qPCR)

To substantiate our findings, we prospectively collected a second, independant set of urine and blood culture samples from two different health care centres: from the University Hospital of Basel between 05/2022 and 06/2022 (= centre 1) and the Institute of Medical Microbiology of the University of Zurich between 09/2022 and 02/2023 (= centre 2).

We screened these samples for the presence of *papGII* carrying *E. coli* using qPCR. Our assay included primers (i) for *E. coli* specific core genes (*gapDH-C* ^66, 67^ and *uidA* ^68^ at centre 1 and *rpoD* at centre 2), (ii) for *papC*, to detect the *pap*-operon and (iii) for *papGII* to specifically detect this variant of *papG*. The sequences of the primers and probes can be found in **Supplementary Methods**. We verified whether there were variants of these sequences in our set of genomes (n=1,076) using the variant caller Freebayes (v1.2.0) ^62^ via snippy (minimum mapping quality=60, minimum basequality=30, minimum coverage=30, minimum proportion for variant evidence=0.95, v4.3.6) (https://github.com/tseemann/snippy). We further tested the performance of our assay when being applied directly to urine sample pellets, thereby decreasing turnaround time by omitting cultivation. We assessed the efficiency and the limit of detection our qPCR assay performed directly on samples and compared them to values resulting from qPCR on extracted genomic DNA.

We used the established qPCR method to prospectively screen clinical urine samples (n=543) at centre 1. This included urine samples collected in routine diagnostics, including 261 samples of 244 cases, which were culture positive for *E. coli*. In contrast at centre 2, we did not perform our qPCR directly from urine samples, but from *E. coli* cultures isolated from urine or from blood culture samples in routine diagnostics (1128 samples from 886 clinical cases). We evaluated which clinical cases progressed to an invasive infection, which we defined as a positive blood culture within 7 days after the retrieval of the urine sample. More details can be found in **Supplementary Methods.**

## Data Availability

The raw reads acquired for this study have been submitted to SRA and are publicly available (Project Accession PRJEB55855). The MALDI-TOF mass spectra acquired for this study can be accessed via the Open Science Foundation (https://osf.io/vmqc5/). All code which was used to visualise bacterial data analysed in this study is available on GitHub (https://github.com/acuenod111/UPEC).

## Acknowledgments

We thank Magdalena Schneider, Christine Kiessling, Elisabeth Schultheiss, Rosa-Maria Vesco, Clarisse Straub, Josiane Reist, Olivia Grüninger, Daniela Lang, and Diana Albertos-Torres for the excellent technical assistance with strain collection, library preparations, and sequencing of the bacterial isolates (all University Hospital Basel). We thank Dr. Deborah R. Vogt (Department of Clinical Research, University of Basel and University Hospital Basel, Basel, Switzerland) for consultations regarding the GLM analysis and Dr. Michael Biggel (University of Zurich) and Dr. Lucas Boeck (University of Basel) for consultation regarding the GWAS analysis. We thank Dr. Gal Horesh (Sanger Institute, UK) and Dr. Thomas Simonet (École Polytechnique Fédérale de Lausanne) for stimulating discussions about *E. coli*, *papG* and valuable feedback on the PCR evaluation as well as Dr. Karoline Leuzinger for consultations regarding qPCR. We thank Dr. Fanny Wegner for valuable feedback on this manuscript. Calculations were performed at sciCORE (http://scicore.unibas.ch/) scientific computing center at University of Basel.

## Author Contributions

Collection and whole genome sequencing of *E. coli* strains: AC, HSS, TR

Collection and curation of clinical data: JA, ST-S, SB, MS, CHN

Quality control of the whole genome sequence data: AC, HSS, TR

Bioinformatic sequence analyses: AC, HSS, TK, TR

MALDI-TOF mass spectra acquisition and analyses: AC, VP

qPCR assay development and validation: AC, DW, DS, AS

Writing of the original manuscript: AC

Providing critical feedback on the manuscript: AC, JA, HSS, TR, DW, ST-S, SB, MS, CHN, JM-G, TK, VP, NRT, AE

Supervision of the project: AE, NRT

Experimental design and study design: AC, AE

## References

1. Öztürk, R. & Murt, A. Epidemiology of urological infections: a global burden. World J Urol 38, 2669–2679 (2020).

2. Mandell, Douglas, and Bennett’s infectious disease essentials. (Elsevier, 2017).

3. Flores-Mireles, A. L., Walker, J. N., Caparon, M. & Hultgren, S. J. Urinary tract infections: epidemiology, mechanisms of infection and treatment options. Nature Reviews Microbiology 13, 269–284 (2015).

4. de Kraker, M. E. A. et al. The changing epidemiology of bacteraemias in Europe: trends from the European Antimicrobial Resistance Surveillance System. Clin Microbiol Infect 19, 860–868 (2013).

5. McNally, A. et al. Diversification of Colonization Factors in a Multidrug-Resistant Escherichia coli Lineage Evolving under Negative Frequency-Dependent Selection. mBio 10, (2019).

6. Arefian, H. et al. Hospital-related cost of sepsis: A systematic review. Journal of Infection 74, 107–117 (2017).

7. Seymour, C. W. et al. Time to Treatment and Mortality during Mandated Emergency Care for Sepsis. New England Journal of Medicine 376, 2235–2244 (2017).

8. Clermont Olivier, Christenson Julia K., Denamur Erick, & Gordon David M. The Clermont Escherichia coli phylo-typing method revisited: improvement of specificity and detection of new phylo-groups. Environmental Microbiology Reports 5, 58–65 (2013).

9. Touchon, M. et al. Phylogenetic background and habitat drive the genetic diversification of Escherichia coli. PLOS Genetics 16, e1008866 (2020).

10. Horesh, G. et al. A comprehensive and high--quality collection of Escherichia coli genomes and their genes. Microbial Genomics 15.

11. Abram, K. et al. Mash-based analyses of Escherichia coli genomes reveal 14 distinct phylogroups. Commun Biol 4, 1–12 (2021).

12. Khairy, R. M., Mohamed, E. S., Ghany, H. M. A. & Abdelrahim, S. S. Phylogenic classification and virulence genes profiles of uropathogenic *E. coli* and diarrhegenic *E. coli* strains isolated from community acquired infections. PLOS ONE 14, e0222441 (2019).

13. Biggel, M. et al. Horizontally acquired *papGII* -containing pathogenicity islands underlie the emergence of invasive uropathogenic Escherichia coli lineages. Nature Communications 11, 5968 (2020).

14. Lloyd, A. L., Smith, S. N., Eaton, K. A. & Mobley, H. L. T. Uropathogenic *Escherichia coli* Suppresses the Host Inflammatory Response via Pathogenicity Island Genes sisA and sisB. Infect Immun 77, 5322–5333 (2009).

15. Mobley, H. L. et al. Isogenic P-fimbrial deletion mutants of pyelonephritogenic *Escherichia coli*: the role of alpha Gal(1-4) beta Gal binding in virulence of a wild-type strain. Mol Microbiol 10, 143–155 (1993).

16. Riley, L. W. Pandemic lineages of extraintestinal pathogenic *Escherichia coli*. Clin Microbiol Infect 20, 380–390 (2014).

17. Denamur, E. et al. Genome wide association study of *Escherichia coli* bloodstream infection isolates identifies genetic determinants for the portal of entry but not fatal outcome. PLoS Genet 18, e1010112 (2022).

18. Klein, R. D. & Hultgren, S. J. Urinary tract infections: microbial pathogenesis, host-pathogen interactions and new treatment strategies. Nat Rev Microbiol 18, 211–226 (2020).

19. Valenza, G. et al. First report of the new emerging global clone ST1193 among clinical isolates of extended-spectrum β-lactamase (ESBL)-producing Escherichia coli from Germany. J Glob Antimicrob Resist 17, 305–308 (2019).

20. cirA - Colicin I receptor - *Escherichia coli* (strain K12) | UniProtKB | UniProt. https://www.uniprot.org/uniprotkb/P17315/entry.

21. hlyA - Hemolysin, chromosomal - *Escherichia coli* | UniProtKB | UniProt. https://www.uniprot.org/uniprotkb/P09983/entry.

22. hlyB - Alpha-hemolysin translocation ATP-binding protein HlyB - *Escherichia coli* | UniProtKB | UniProt. https://www.uniprot.org/uniprotkb/P08716/entry.

23. hlyD - Hemolysin secretion protein D, chromosomal - *Escherichia coli* | UniProtKB | UniProt. https://www.uniprot.org/uniprotkb/P09986/entry.

24. cbtA - Cytoskeleton-binding toxin CbtA - *Escherichia coli* (strain K12) | UniProtKB | UniProt. https://www.uniprot.org/uniprotkb/P64524/entry.

25. cbeA - Cytoskeleton bundling-enhancing antitoxin CbeA - *Escherichia coli* (strain K12) | UniProtKB | UniProt. https://www.uniprot.org/uniprotkb/P76364/entry.

26. Fagerquist, C. K. et al. Rapid identification of protein biomarkers of *Escherichia coli* O157:H7 by matrix-assisted laser desorption ionization-time-of-flight-time-of-flight mass spectrometry and top-down proteomics. Anal Chem 82, 2717–2725 (2010).

27. Holt, K. E., Lassalle, F., Wyres, K. L., Wick, R. & Mostowy, R. J. Diversity and evolution of surface polysaccharide synthesis loci in Enterobacteriales. ISME J 14, 1713–1730 (2020).

28. Matsuura, M. Structural Modifications of Bacterial Lipopolysaccharide that Facilitate Gram-Negative Bacteria Evasion of Host Innate Immunity. Frontiers in Immunology 4, 109 (2013).

29. Goh, K. G. K. et al. Genome-Wide Discovery of Genes Required for Capsule Production by Uropathogenic Escherichia coli. mBio 8, e01558–17.

30. McNally, A. et al. Genomic analysis of extra-intestinal pathogenic Escherichia coli urosepsis. Clinical Microbiology and Infection 19, e328–e334 (2013).

31. Levert, M. et al. Molecular and Evolutionary Bases of Within-Patient Genotypic and Phenotypic Diversity in Escherichia coli Extraintestinal Infections. PLOS Pathogens 6, e1001125 (2010).

32. Strömberg, N. et al. Host-specificity of uropathogenic *Escherichia coli* depends on differences in binding specificity to Gal alpha 1-4Gal-containing isoreceptors. EMBO J 9, 2001–2010 (1990).

33. Legros, N. et al. PapG subtype-specific binding characteristics of *Escherichia coli* towards globo-series glycosphingolipids of human kidney and bladder uroepithelial cells. Glycobiology 29, 789–802 (2019).

34. Dodson, K. W. et al. Structural basis of the interaction of the pyelonephritic *E. coli* adhesin to its human kidney receptor. Cell 105, 733–743 (2001).

35. Ambite, I. et al. Fimbriae reprogram host gene expression – Divergent effects of P and type 1 fimbriae. PLoS Pathog 15, e1007671 (2019).

36. Urinary Tract Infections | Mandell, Douglas, and Bennett’s Principles…. https://expertconsult.inkling.com/read/mandell-douglas-bennetts-infectious-diseases-8/chapter-74/urinary-tract-infections.

37. Dodson, K. W. et al. Structural Basis of the Interaction of the Pyelonephritic *E. coli* Adhesin to Its Human Kidney Receptor. Cell 105, 733–743 (2001).

38. Ambite, I. et al. Fimbriae reprogram host gene expression – Divergent effects of P and type 1 fimbriae. PLOS Pathogens 15, e1007671 (2019).

39. https://www.uniprot.org/uniprot/Q47450. *Uniprot, papGII*.

40. Sauget, M., Valot, B., Bertrand, X. & Hocquet, D. Can MALDI-TOF Mass Spectrometry Reasonably Type Bacteria? Trends in Microbiology 25, 447–455 (2017).

41. Gupta, K., Grigoryan, L. & Trautner, B. Urinary Tract Infection. Ann Intern Med 167, ITC49–ITC64 (2017).

42. Charlson, M., Szatrowski, T. P., Peterson, J. & Gold, J. Validation of a combined comorbidity index. J Clin Epidemiol 47, 1245–1251 (1994).

43. Bolger, A. M., Lohse, M. & Usadel, B. Trimmomatic: a flexible trimmer for Illumina sequence data. Bioinformatics 30, 2114–2120 (2014).

44. Bankevich, A. et al. SPAdes: a new genome assembly algorithm and its applications to single-cell sequencing. J Comput Biol 19, 455–477 (2012).

45. Wick, R. R., Judd, L. M., Gorrie, C. L. & Holt, K. E. Unicycler: Resolving bacterial genome assemblies from short and long sequencing reads. PLOS Computational Biology 13, e1005595 (2017).

46. Walker, B. J. et al. Pilon: An Integrated Tool for Comprehensive Microbial Variant Detection and Genome Assembly Improvement. PLOS ONE 9, e112963 (2014).

47. Truong, D. T. et al. MetaPhlAn2 for enhanced metagenomic taxonomic profiling. Nat Methods 12, 902–903 (2015).

48. Seemann, T. Prokka: rapid prokaryotic genome annotation. Bioinformatics 30, 2068– 2069 (2014).

49. Jolley, K. A. et al. Ribosomal multilocus sequence typing: universal characterization of bacteria from domain to strain. Microbiology 158, 1005–1015 (2012).

50. Feldgarden, M. et al. AMRFinderPlus and the Reference Gene Catalog facilitate examination of the genomic links among antimicrobial resistance, stress response, and virulence. Sci Rep 11, 12728 (2021).

51. Ondov, B. D. et al. Mash: fast genome and metagenome distance estimation using MinHash. Genome Biology 17, 132 (2016).

52. Abram, K. et al. Mash-based analyses of Escherichia coli genomes reveal 14 distinct phylogroups. Commun Biol 4, 117 (2021).

53. Inouye, M. et al. SRST2: Rapid genomic surveillance for public health and hospital microbiology labs. Genome Medicine 6, 90 (2014).

54. Ingle, D. J. et al. In silico serotyping of E. coli from short read data identifies limited novel O-loci but extensive diversity of O:H serotype combinations within and between pathogenic lineages. Microb Genom 2, e000064 (2016).

55. Jain, C., Rodriguez-R, L. M., Phillippy, A. M., Konstantinidis, K. T. & Aluru, S. High throughput ANI analysis of 90K prokaryotic genomes reveals clear species boundaries. Nat Commun 9, 5114 (2018).

56. Tonkin-Hill, G. et al. Producing polished prokaryotic pangenomes with the Panaroo pipeline. Genome Biology 21, 180 (2020).

57. Katoh, K., Misawa, K., Kuma, K. & Miyata, T. MAFFT: a novel method for rapid multiple sequence alignment based on fast Fourier transform. Nucleic Acids Res 30, 3059–3066 (2002).

58. Stamatakis, A. RAxML version 8: a tool for phylogenetic analysis and post-analysis of large phylogenies. Bioinformatics 30, 1312–1313 (2014).

59. Madeira, F. et al. Search and sequence analysis tools services from EMBL-EBI in 2022. Nucleic Acids Res 50, W276–279 (2022).

60. hdeA - Acid stress chaperone HdeA - Escherichia coli (strain K12) | UniProtKB | UniProt. https://www.uniprot.org/uniprotkb/P0AES9/entry.

61. Gasteiger, E. et al. Protein Identification and Analysis Tools on the ExPASy Server. in The Proteomics Protocols Handbook (ed. Walker, J. M.) 571–607 (Humana Press, 2005). doi:10.1385/1-59259-890-0:571.

62. Garrison, E. & Marth, G. Haplotype-based variant detection from short-read sequencing. Preprint at https://doi.org/10.48550/arXiv.1207.3907 (2012).

63. Lees, J. A., Galardini, M., Bentley, S. D., Weiser, J. N. & Corander, J. pyseer: a comprehensive tool for microbial pangenome-wide association studies. Bioinformatics 34, 4310–4312 (2018).

64. Jaillard, M. et al. A fast and agnostic method for bacterial genome-wide association studies: Bridging the gap between k-mers and genetic events. PLoS Genet 14, e1007758 (2018).

65. Gibb, S. & Strimmer, K. MALDIquant: a versatile R package for the analysis of mass spectrometry data. Bioinformatics 28, 2270–2271 (2012).

66. gapC - Glyceraldehyde-3-phosphate dehydrogenase C - Escherichia coli O157:H7 | UniProtKB | UniProt. https://www.uniprot.org/uniprotkb/P58072/entry.

67. Hidalgo, E., Limón, A. & Aguilar, J. A second Escherichia coli gene with similarity to gapA. Microbiologia 12, 99–106 (1996).

68. Bej, A. K., DiCesare, J. L., Haff, L. & Atlas, R. M. Detection of Escherichia coli and Shigella spp. in water by using the polymerase chain reaction and gene probes for uid. Appl Environ Microbiol 57, 1013–1017 (1991).

