## Supplementary material for "Bacterial genome wide association study substantiates *papGII* of *Escherichia coli* as a patient independent driver of urosepsis": Figure_S1.pdf

### Gender

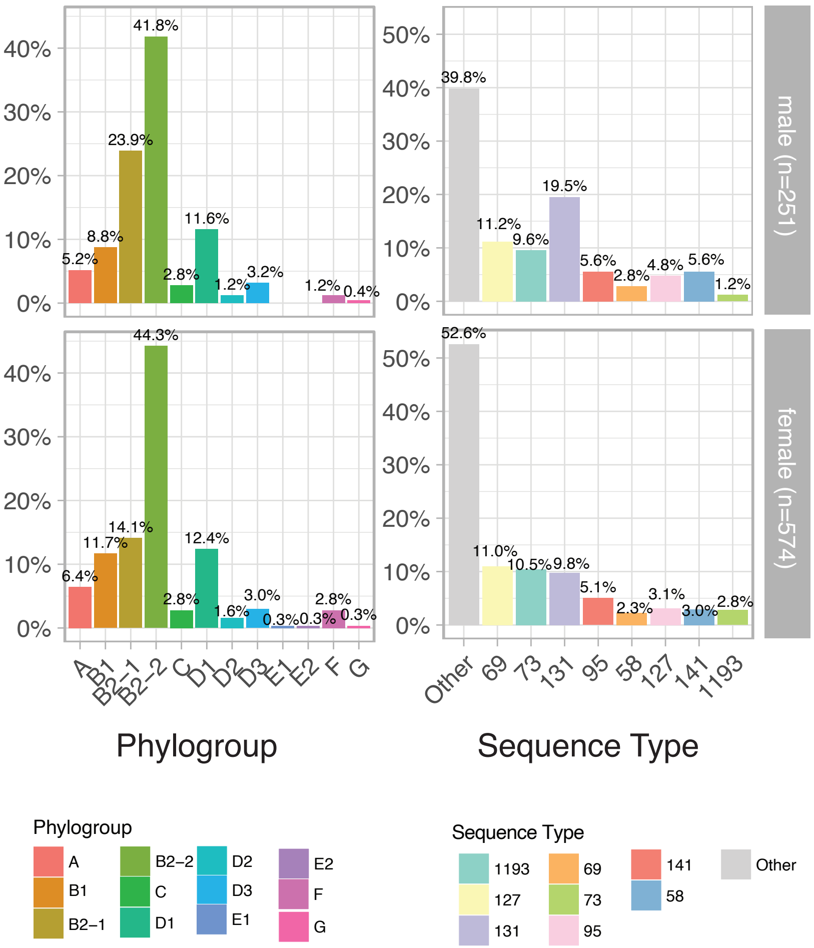


**Figure S1:** Distribution of *E. coli* phylogroups (left) and Sequence Types (ST) (right) in male (n=251) (upper row) and female (n=574) (lower row) patients.
