## Supplementary material for "Bacterial genome wide association study substantiates *papGII* of *Escherichia coli* as a patient independent driver of urosepsis": Figure_S2.pdf

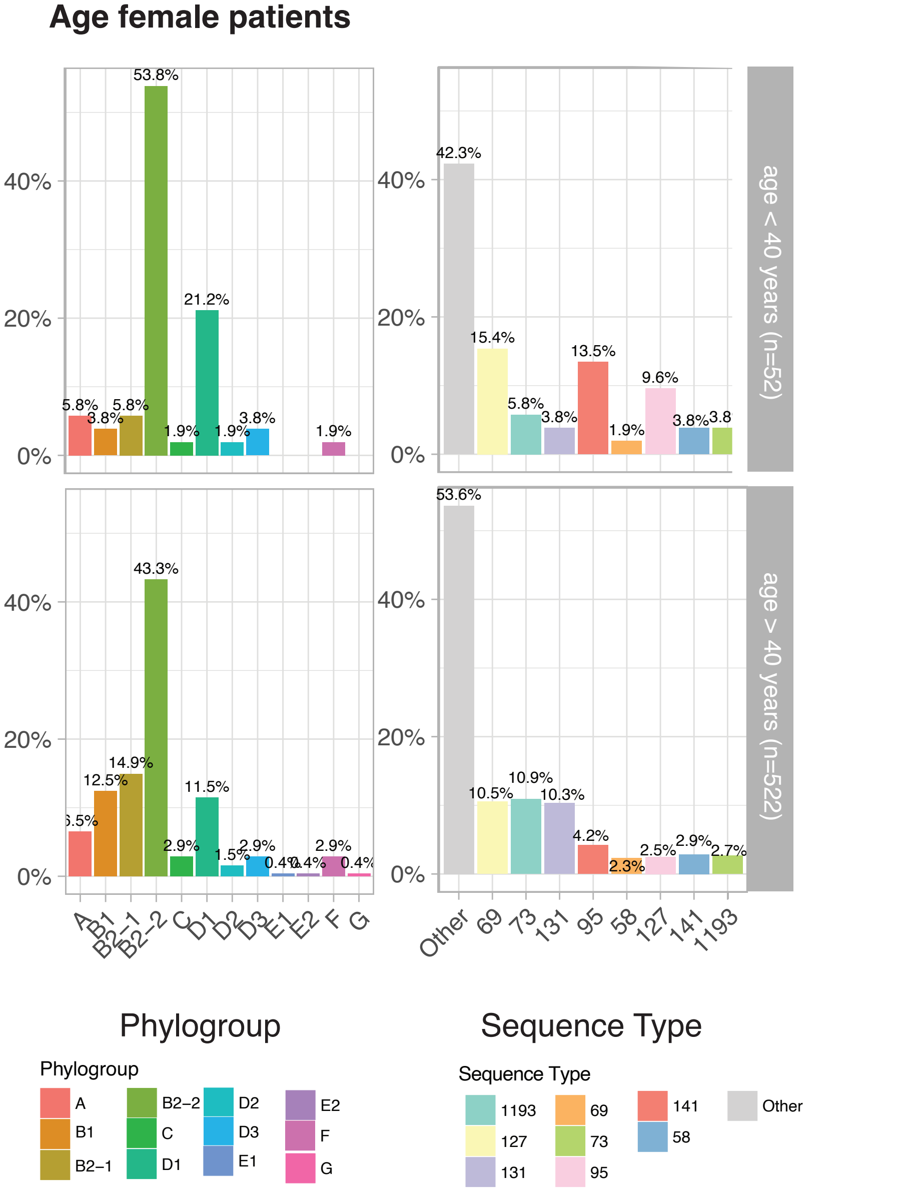


**Figure S2:** Distribution of *E. coli* phylogroups (left) and Sequence Types (ST) (right) in in female patients (n=574) younger than 40 years (n=52) (upper row) and older than 40 years (n=522) (lower row) patients.
