## Supplementary material for "Bacterial genome wide association study substantiates *papGII* of *Escherichia coli* as a patient independent driver of urosepsis": Figure_S3.pdf

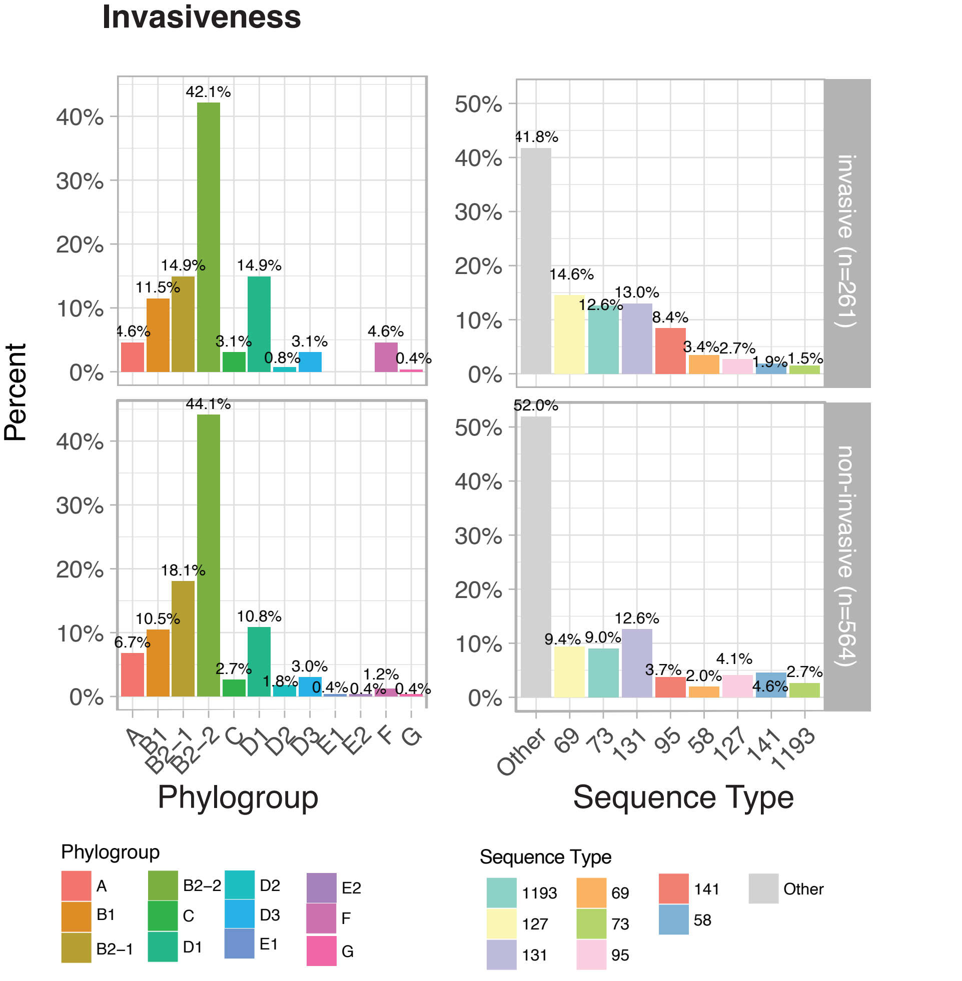


**Figure S3:** Distribution of *E. coli* phylogroups (left) and Sequence Types (ST) (right) in invasive infections (n=261) (upper row) and non-invasive infections (n=574) (lower row) patients.
