## Supplementary material for "Bacterial genome wide association study substantiates *papGII* of *Escherichia coli* as a patient independent driver of urosepsis": Figure_S4.pdf

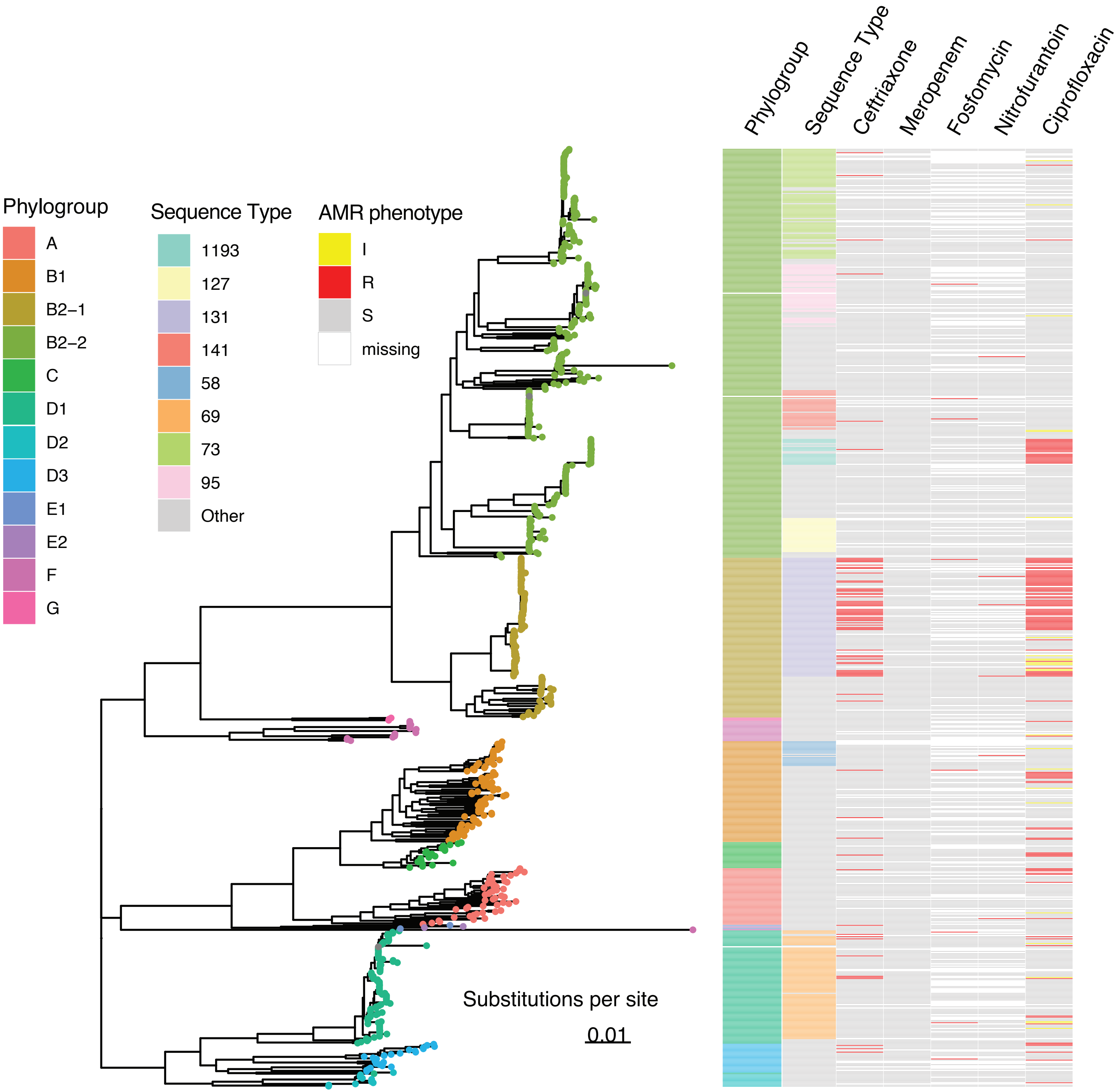


**Figure S4:** Core genome phylogeny of 825 *E. coli* strains. Columns represent (from left to right): the assigned phylogroup, the sequence type, phenotypic resistance against Ceftriaxone, Meropenem, Fosfomycin, Nitrofurantoin and Ciprofloxacin.
