## Supplementary material for "Bacterial genome wide association study substantiates *papGII* of *Escherichia coli* as a patient independent driver of urosepsis": Figure_S5.pdf

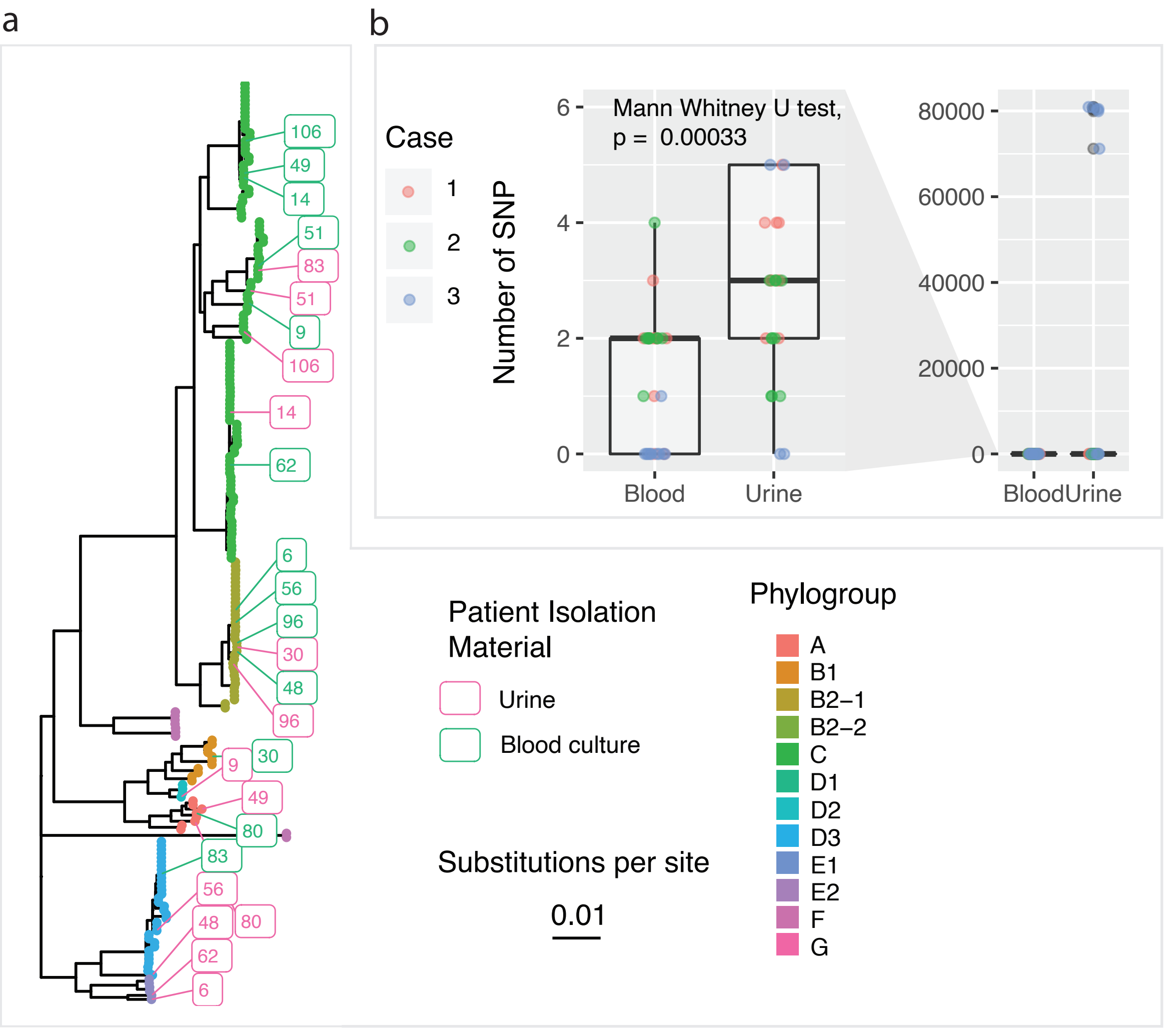


**Figure S5:** Within host genetic diversity of E. coli strains isolated from the same clinical cases **a:** core genome phylogeny of E. coli strains (n=225), isolated from the same clinical case (n=106), colored by phylogroup. The numbers correspond to the case identifier and strains were only labelled, if they exhibited < 99.9% Average Nucleotide Identity to the strain isolated from the same clinical case **b:** SNV for of of 10 picked isolates from three cases, either from urine or blood culture samples.
