## Supplementary material for "Bacterial genome wide association study substantiates *papGII* of *Escherichia coli* as a patient independent driver of urosepsis": Figure_S6.pdf

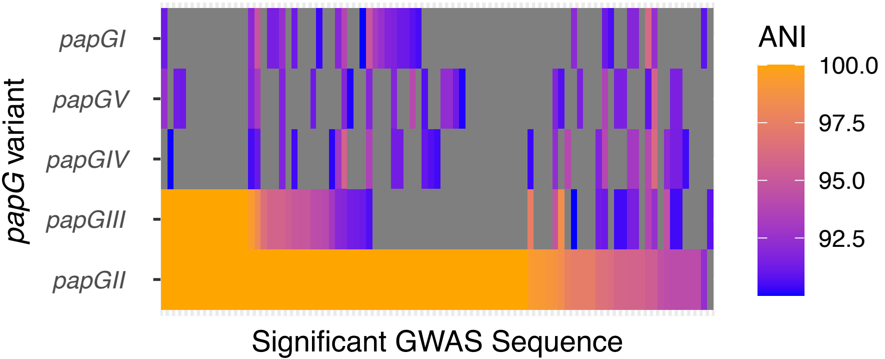


**Figure S6:** Average Nucleotide identity values for unitigs identified in our bGWAS and mapping to *papG* (X-axis) and the reference sequences for the five *papG* variants (Y-axis).
