## Supplementary material for "Bacterial genome wide association study substantiates *papGII* of *Escherichia coli* as a patient independent driver of urosepsis": Figure_S7.pdf

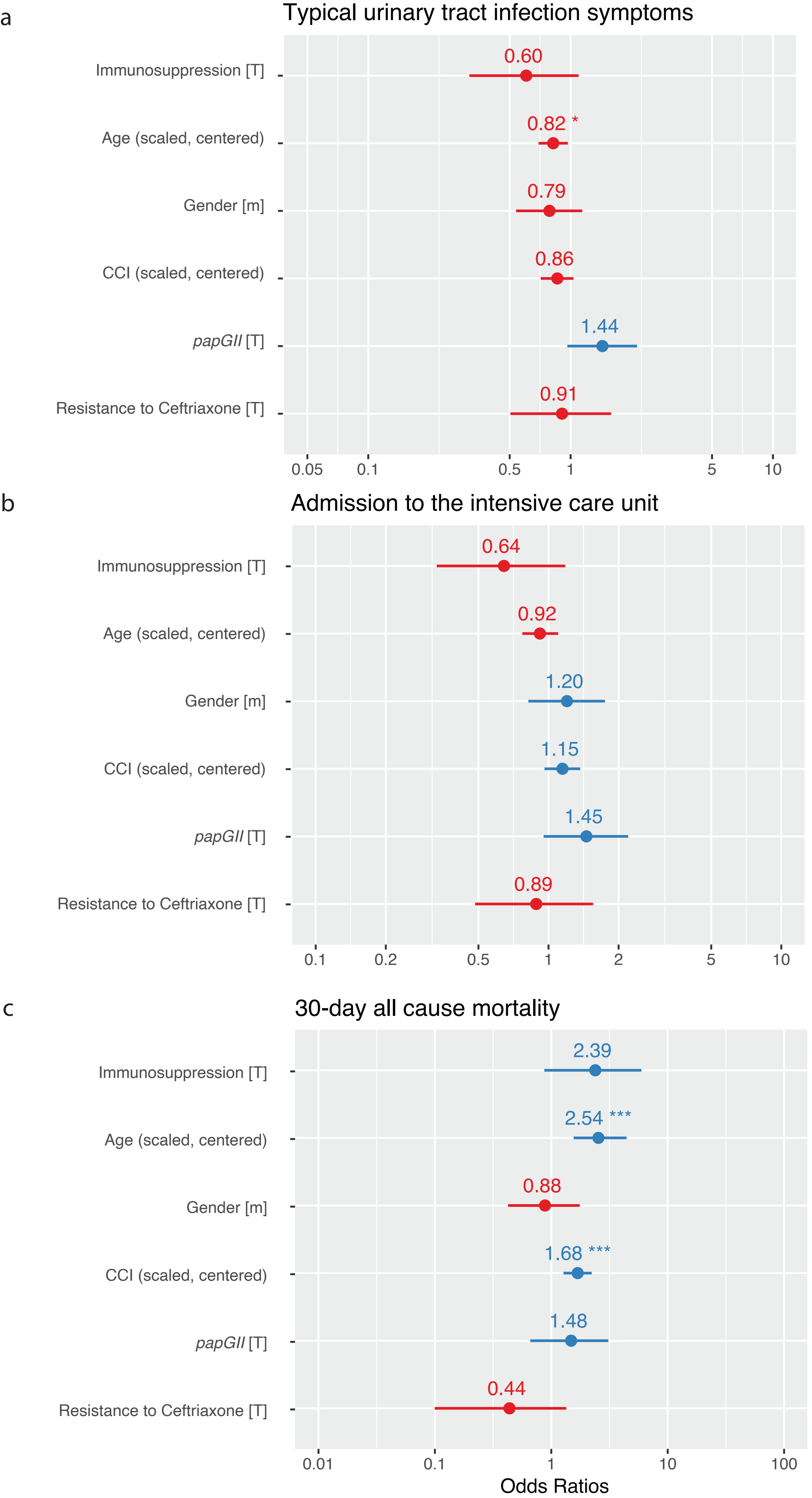


**Figure S7:** Odds ratio estimates with 95% confidence intervals for **a**: Typical urinary tract infection symptoms (n = 717 complete observations with 213 events); **b**: Admission to the intensive care unit (n = 751 complete observations with 172 events); **c**: 30-day all cause mortality (n = 749 complete observations with 45 events); using the generalised linear model (GLM). OR = odds ratio; CI = confidence interval; CCI = Charlson Comorbidity Index.
