## Supplementary material for "Bacterial genome wide association study substantiates *papGII* of *Escherichia coli* as a patient independent driver of urosepsis": Figure_S8.pdf

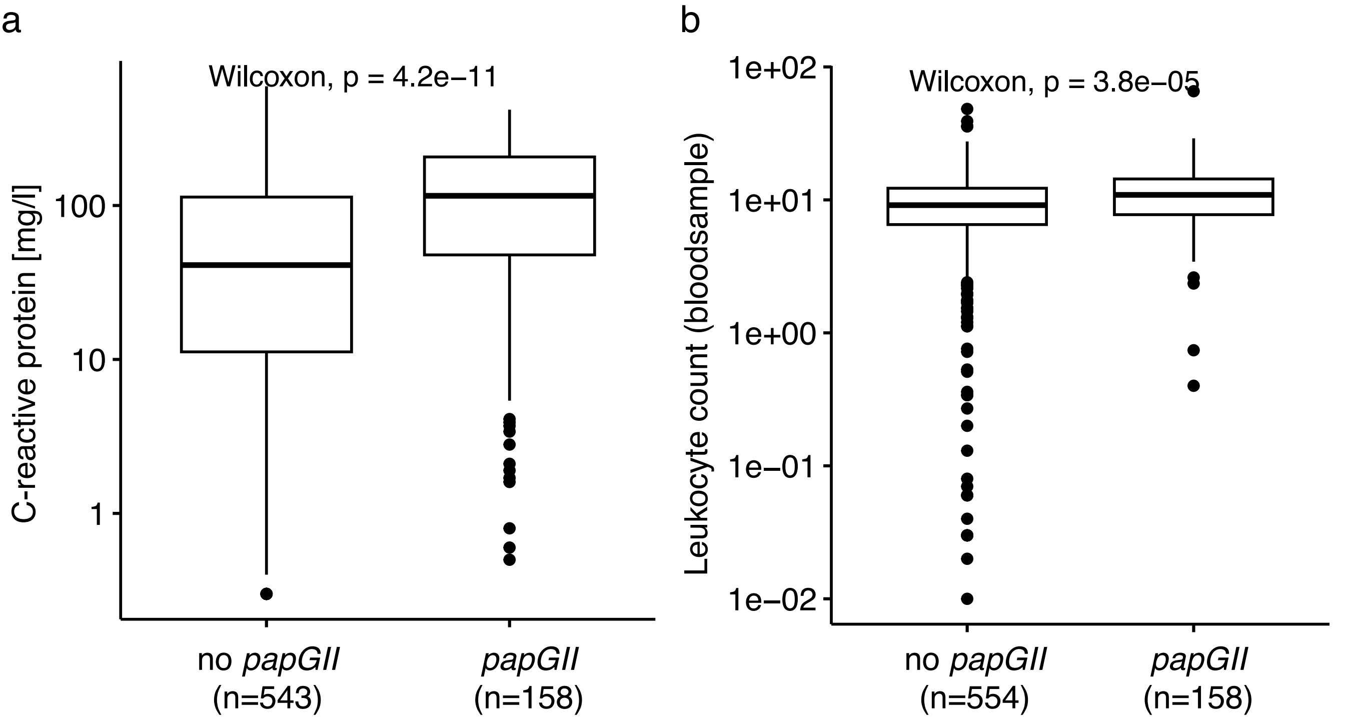


**Figure S8:** C-reactive protein concentration (**a**) and leucocyte count (**b**) measured in blood samples of cases, for which a *papGII* positive or a *papGII* negative *E. coli* strain was isolated from a urine or a blood culture samples. Leucocyte counts were measured on the day the urine / bloodculture samples were taken.
