## Supplementary material for "Bacterial genome wide association study substantiates *papGII* of *Escherichia coli* as a patient independent driver of urosepsis": Figure_S9.pdf

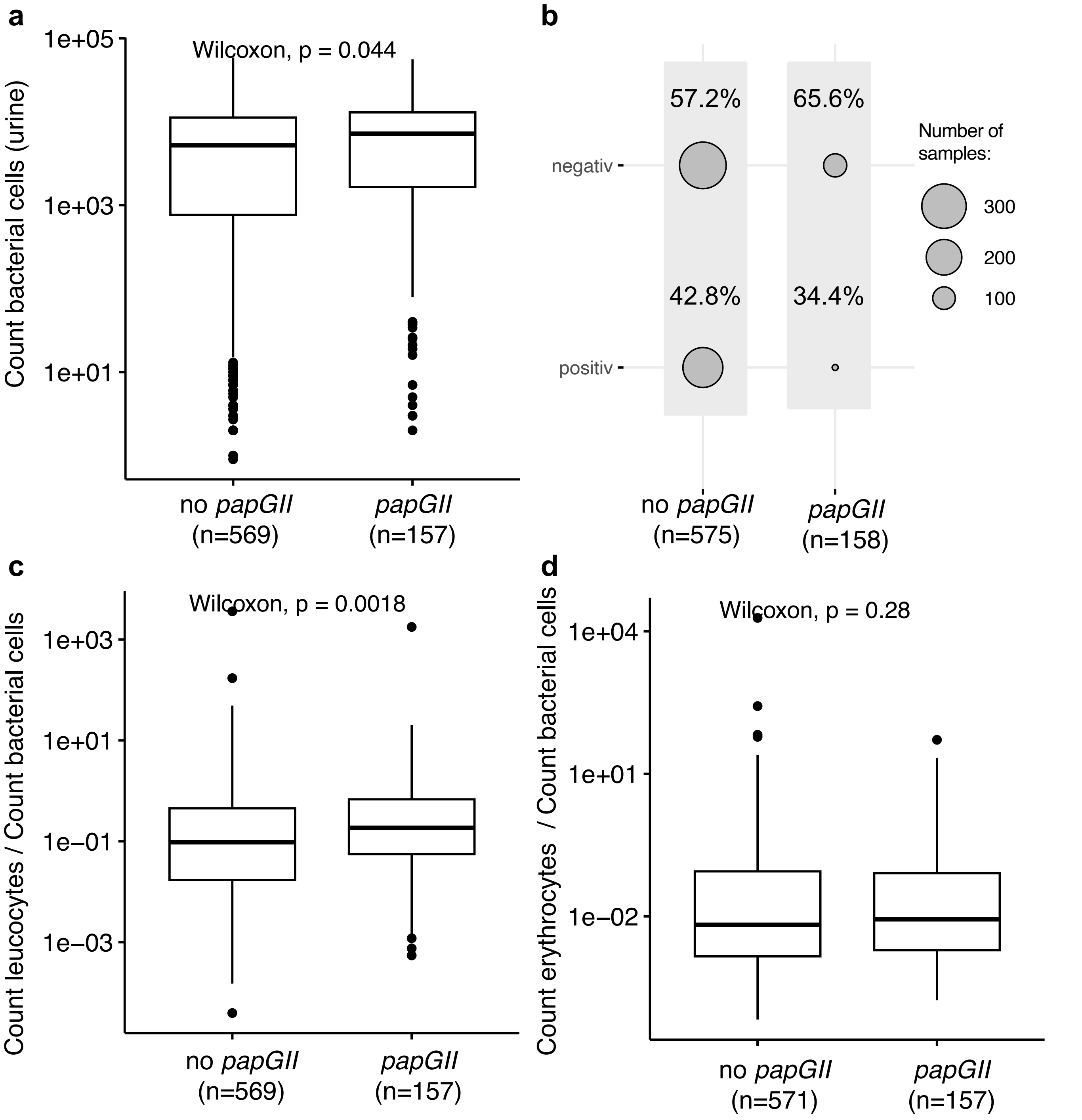


**Figure S9:** Bacterial cell count (**a**), nitrite status (**b**), leucocyte count divided by bacterial cell count (**c**) and erythrocyte count divided by bacterial cell count (**d**) measured in urine samples of cases, for which a *papGII* positive or a *papGII* negative *E. coli* strain was isolated from a urine or a blood culture samples.
