## Supplementary material for "Bacterial genome wide association study substantiates *papGII* of *Escherichia coli* as a patient independent driver of urosepsis": Figure_S10.pdf

### *papGII* occurrence

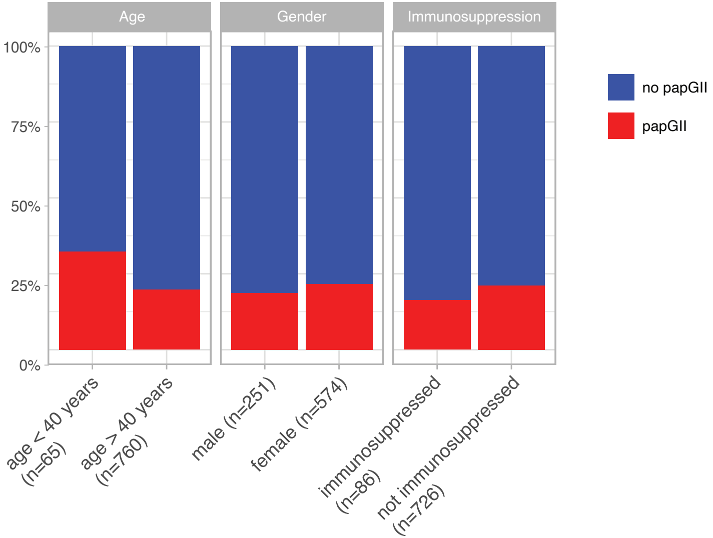


**Figure S10:** Relative occurrence of *papGII* in isolates from patients younger vs. older than 40 years, in isolates from male vs. female patients and in isolates from patients which were immunosuppressed vs. patients which were not immunosuppressed.
