## Supplementary material for "Bacterial genome wide association study substantiates *papGII* of *Escherichia coli* as a patient independent driver of urosepsis": Figure_S11.pdf

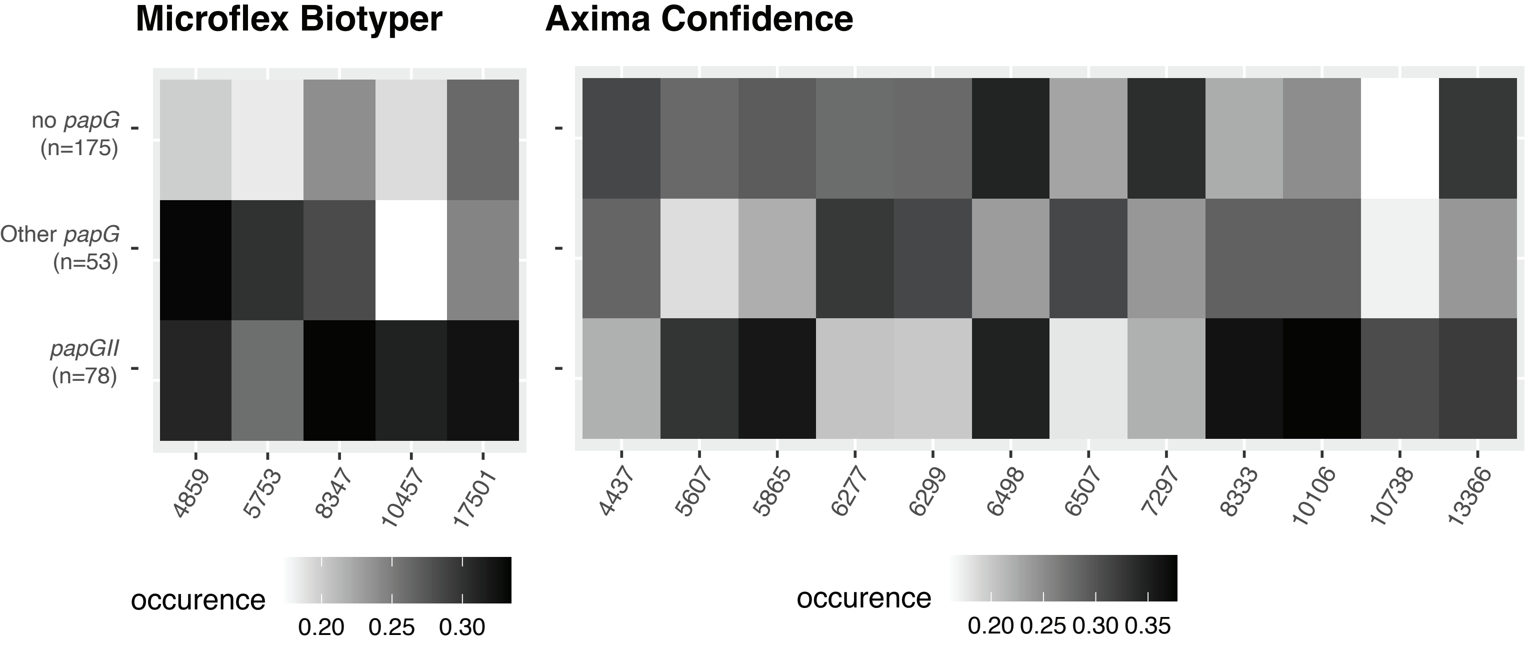


**Figure S11:** Occurrence of MALDI-TOF mass peaks in spectra acquired from *E. coli* strains encoding no *papG* gene, encoding a *papG* variant other than papGII and encoding *papGII*. Each strain was measured in quadruplicate either on a Microflex Biotyper device, or an Axmina Confidence device. Masses are only depicted if detected in > 30% or < 25% of spectra for one or more of the groups.
