## Supplementary material for "Bacterial genome wide association study substantiates *papGII* of *Escherichia coli* as a patient independent driver of urosepsis": Figure_S12.pdf

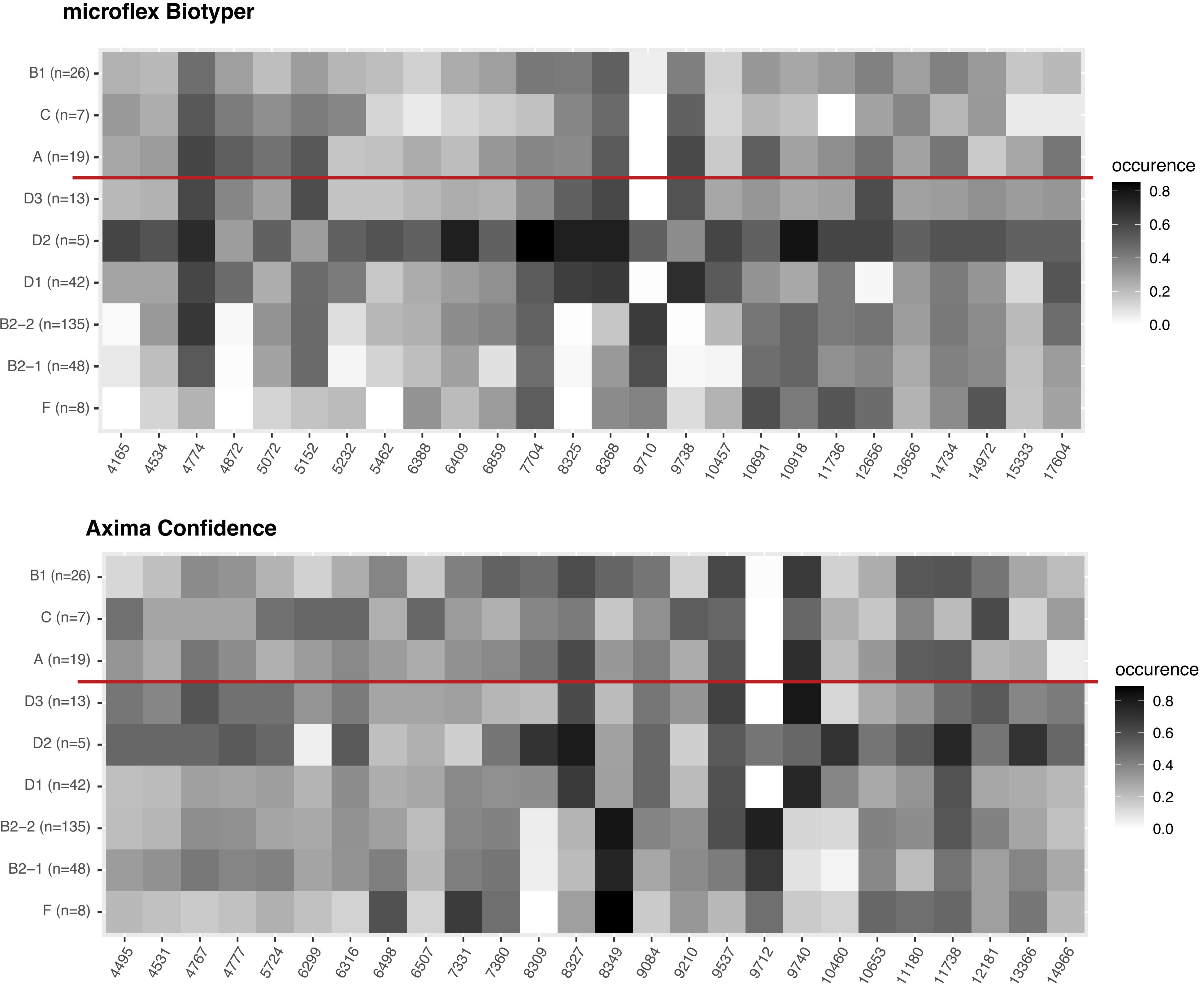


**Figure S12:** Occurrence of MALDI-TOF mass peaks in spectra acquired from *E. coli* strains of different phylogroups. Each strain was measured in quadruplicate either on a Microflex Biotyper device, or an Axmina Confidence device. Phylogroups for which less than five strains were available (E1, E2 and G) were excluded from the plot. Masses are only depicted if detected in > 50% or < 25% of spectra for one or more of the groups.
