## Supplementary material for "Bacterial genome wide association study substantiates *papGII* of *Escherichia coli* as a patient independent driver of urosepsis": Figure_S13.pdf

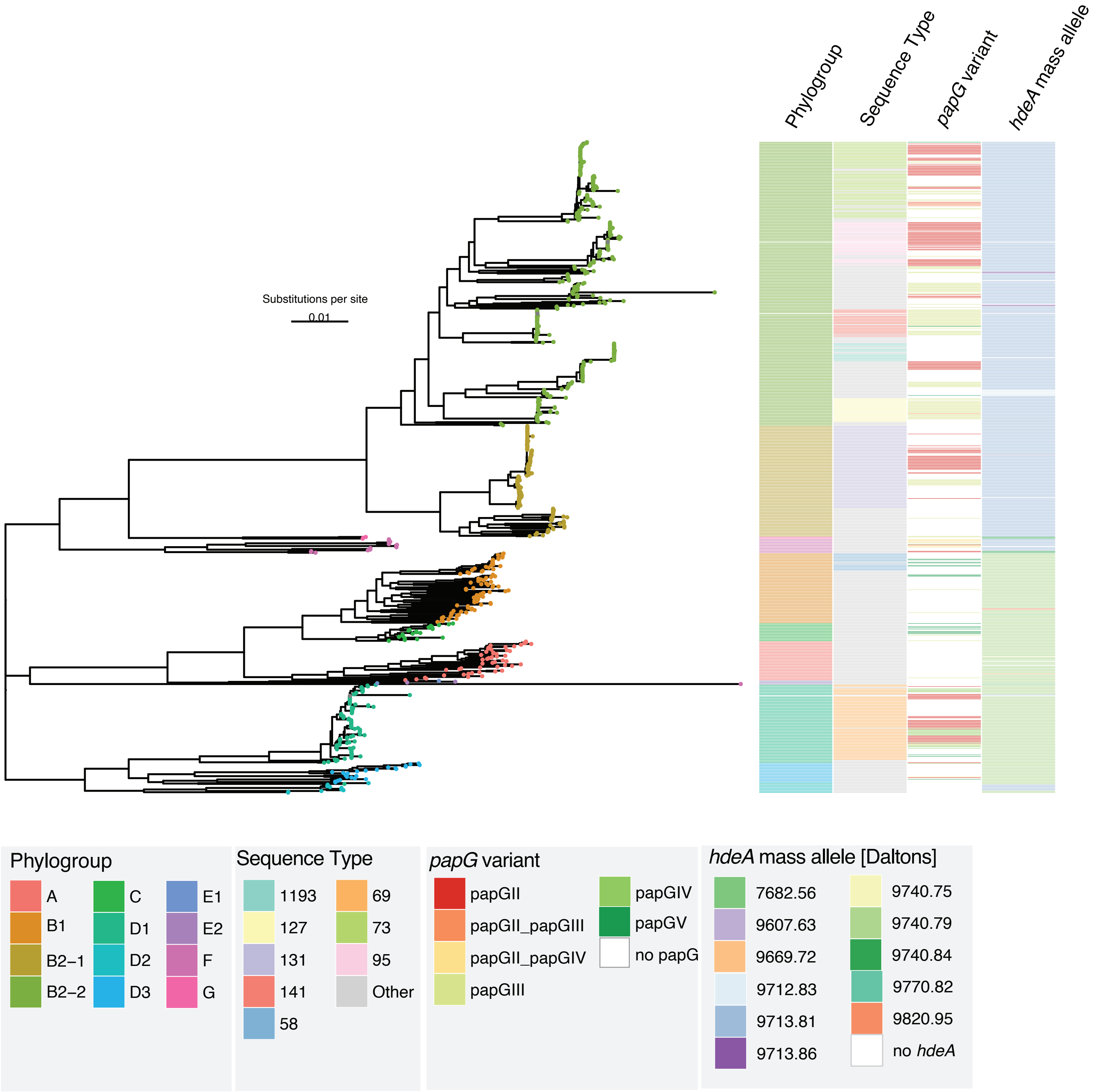


**Figure S13:** Core genome phylogeny of the *E. coli* strains collected for this study (one strain per clinical case, n=825). Phylogroup assignment, Sequence Type (ST) (eight most frequent ones coloured, more rare STs in grey), *papG* variant, mass of HdeA, predicted from the amino acid sequence.
