## Supplementary material for "Bacterial genome wide association study substantiates *papGII* of *Escherichia coli* as a patient independent driver of urosepsis": Figure_S14.pdf

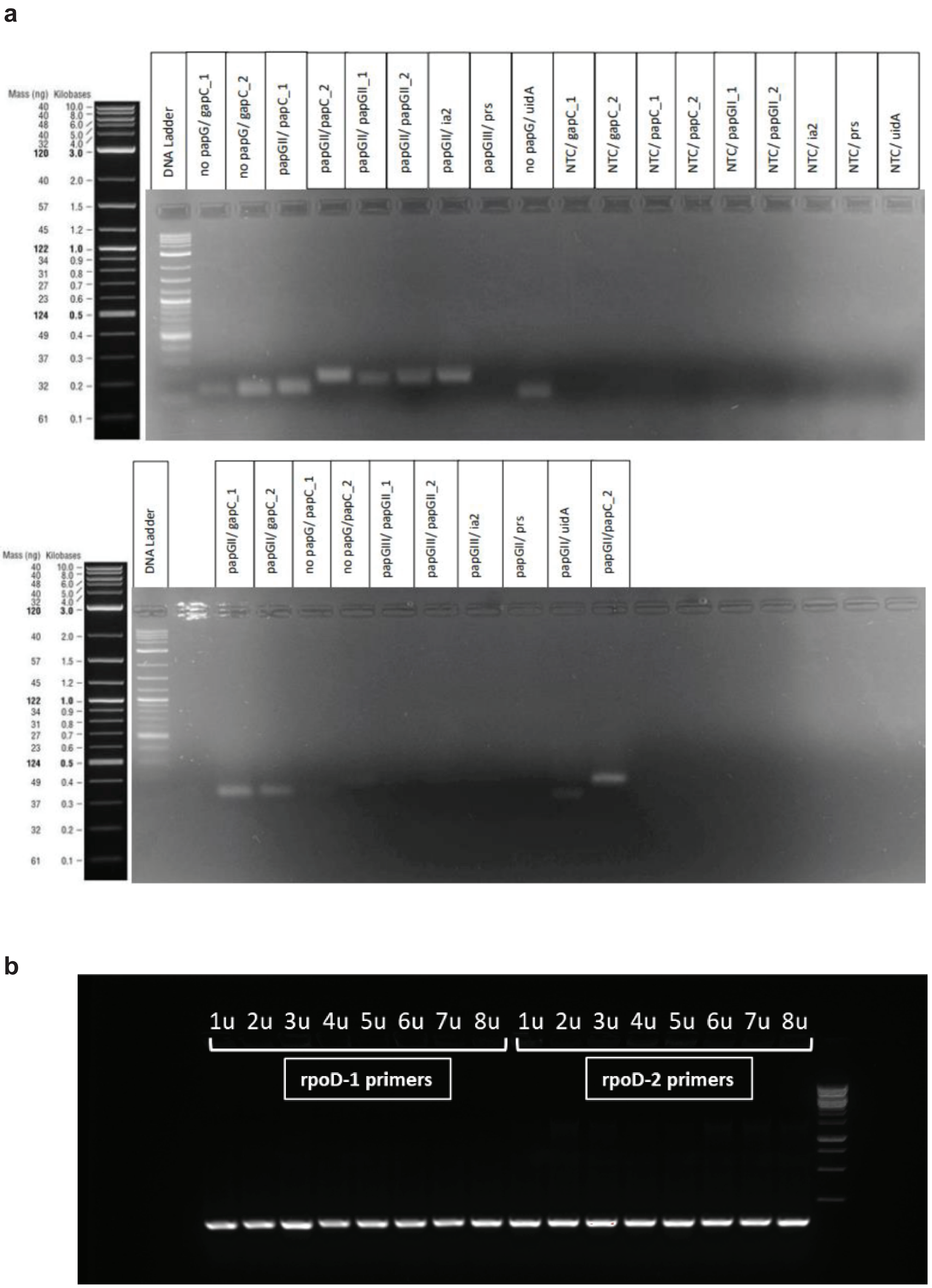


**Figure S14:** Results of the endpoint PCR assay (a) to test the functionality of the primers designed at centre 1. This also includes tests for the crossreactivity between papGII and papGIII primers. (b) to test the functionality of the *rpoD* primers designed at centre 2.
