## Supplementary material for "Bacterial genome wide association study substantiates *papGII* of *Escherichia coli* as a patient independent driver of urosepsis": Figure_S15.pdf

a

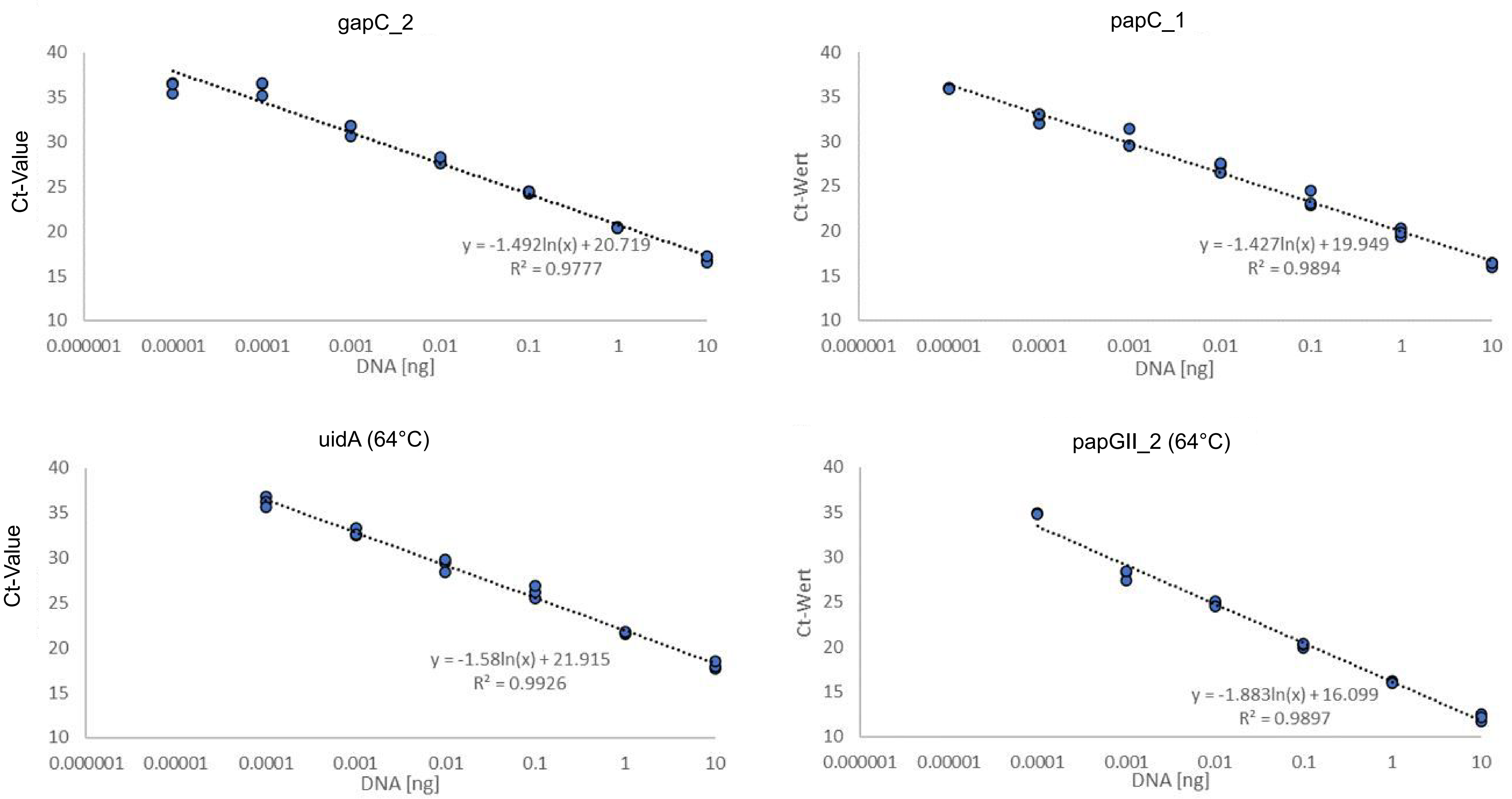


b

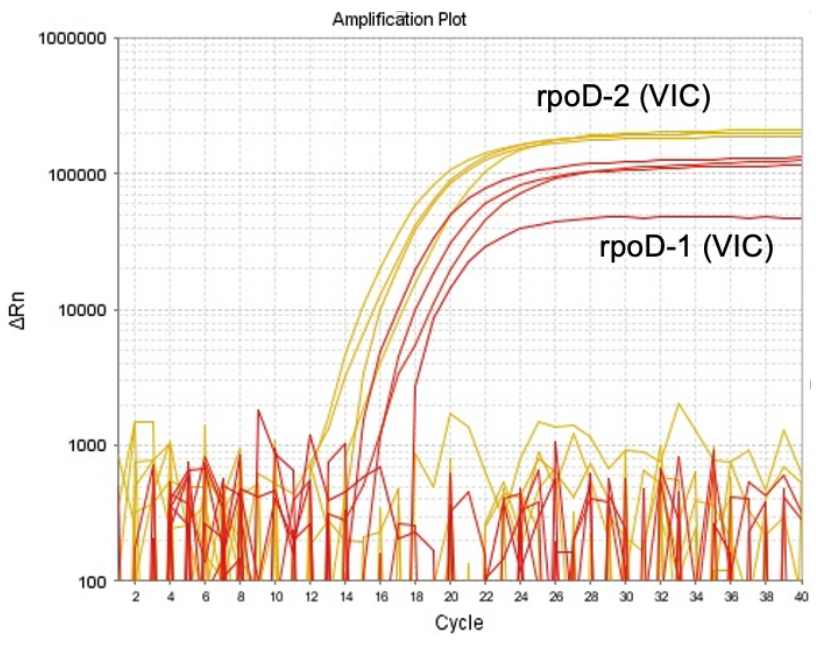


**Figure S15:** Evaluating the efficiency of primers and probes used in our qPCR assay (**a**) qPCR standard curves and values for the primer pairs gapC_2, papC_1, uidA and papGII_2 tested at centre 1. Each measurement was performed in triplicate. (**b**) Amplification plots for the two rpoD probes designed at centre 2. Measurements performed in quadruplicate.
