## Supplementary material for "Bacterial genome wide association study substantiates *papGII* of *Escherichia coli* as a patient independent driver of urosepsis": Figure_S16.pdf

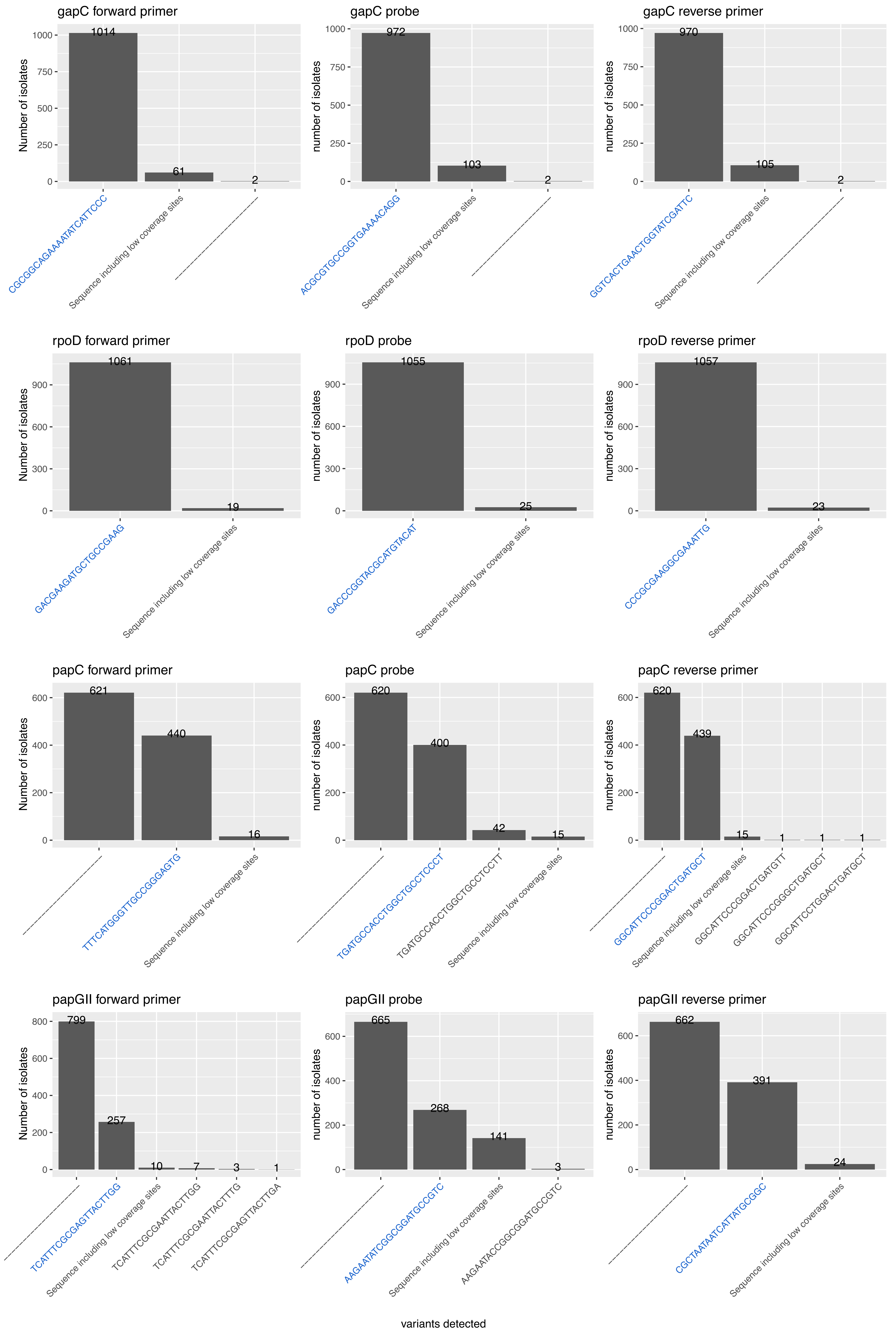


**Figure S16:** Variants of primer and probe sequences detected in our genome collection (n=1,076). Sequences used in the qPCR assay are indicated in blue. Variants were called using the variantcaller Freebayes via snippy and using a minimum coverage of 20x.
