## Supplementary material for "Bacterial genome wide association study substantiates *papGII* of *Escherichia coli* as a patient independent driver of urosepsis": Figure_S17.pdf

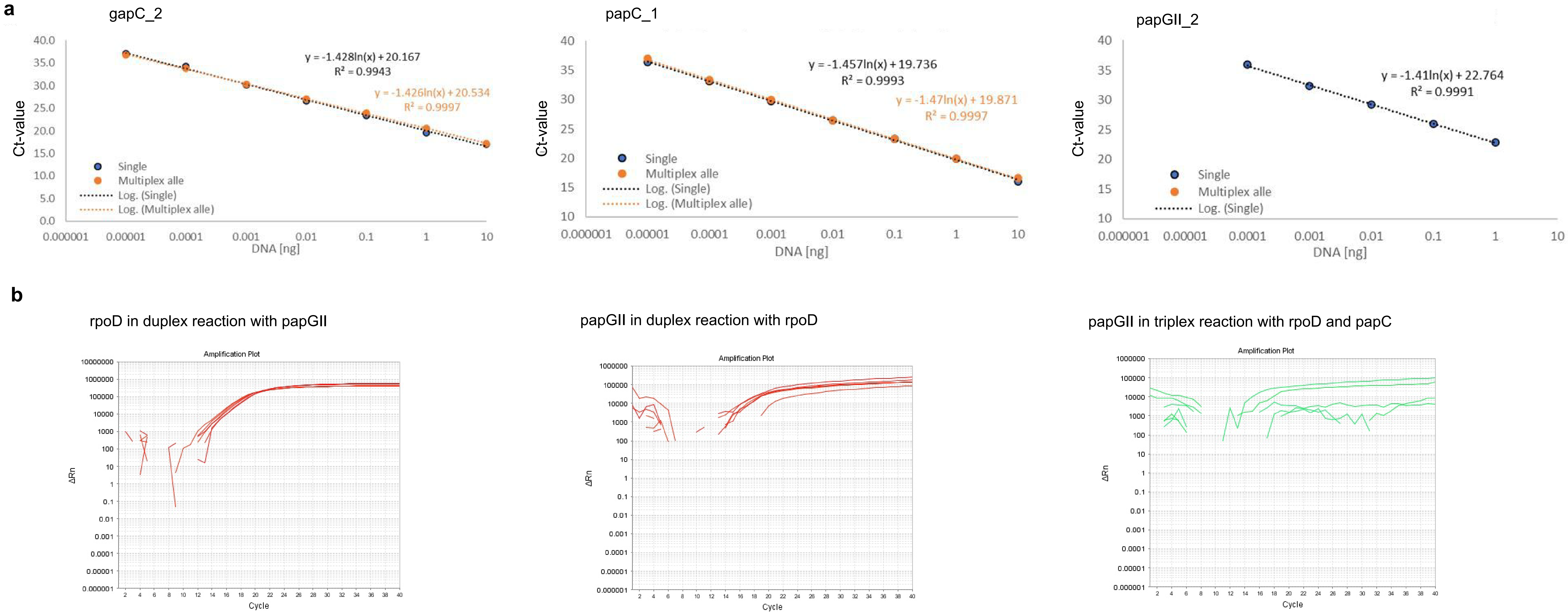


**Figure S17: (a)** Efficiency of the primer pairs in the single reaction (blue) and in a triplex reaction (orange) for the primers used at center 1 (gapC, papC and papGII). **(b)** Amplification curves of primers used at center 2: rpoD and papGII in duplex reactions and of papGII in a triplex reaction with rpoD and papC.
