## Supplementary material for "Bacterial genome wide association study substantiates *papGII* of *Escherichia coli* as a patient independent driver of urosepsis": Figure_S18.pdf

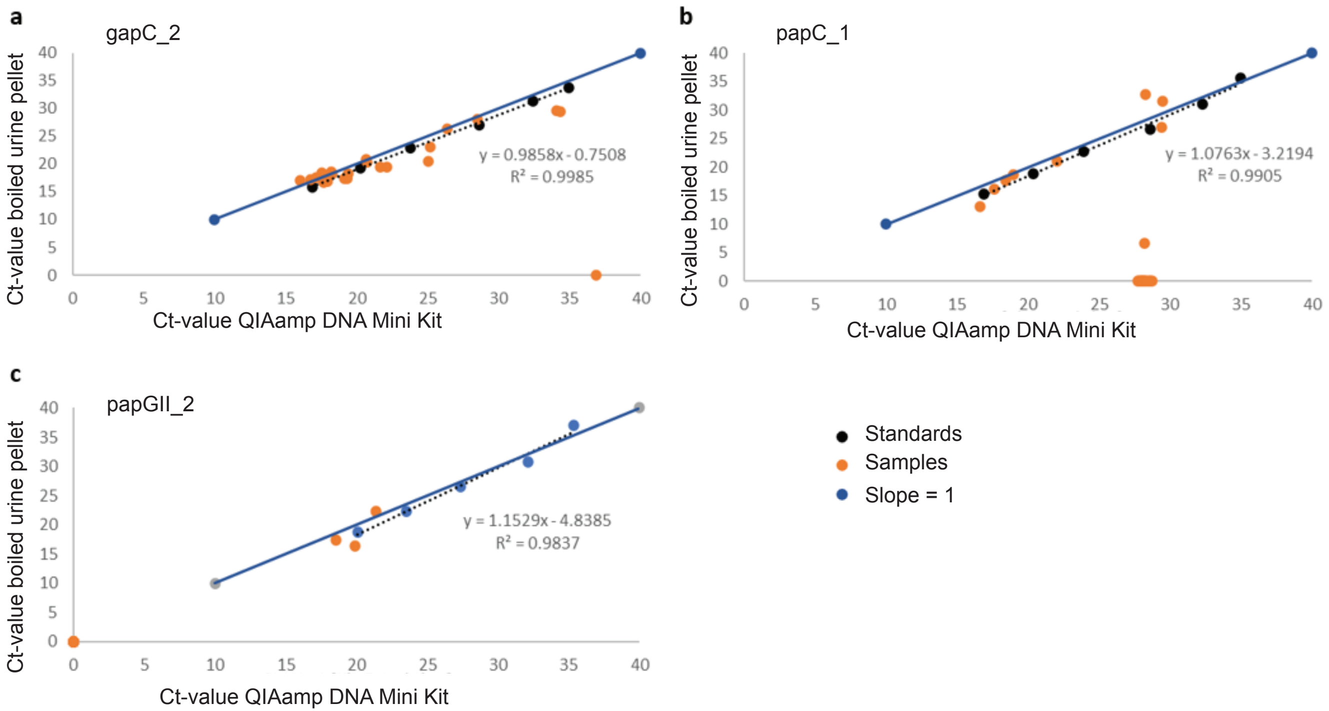


**Figure S18**: Comparison of the Ct-value yielded when processing urine pellets (n=24) using the QIAamp DNA Mini Kit and after boiling for 10 minutes.
