## Supplementary material for "Bacterial genome wide association study substantiates *papGII* of *Escherichia coli* as a patient independent driver of urosepsis": Supplementary Methods.pdf

### Supplementary Methods Polymerase Chain Reaction (PCR)

#### Endpoint PCR

##### Centre 1

To test the functionality of the selected primers we performed an endpoint PCR. The primers used are summarised in **Table 1**. Six primer pairs were newly designed for this study using Primer-BLAST (4) and three were retrieved from previous publications (1,2).

**Table 1:** Primers which were used in the endpoint PCR assay at centre 1. Primers with an asterisk (*) were newly designed for this study.

| Name | Target | Amplified [bp] | Temp. [°C] | Sequence (5‘-3‘) |
|---|---|---|---|---|
| gapC_1* | *gapDH-C* | 140 | 64 | F: TGA TTC AAA CTA TGG TCC GTT CCC<br>R: CAA TGA TTT CTG CAC CTT TCG C |
| gapC_2* | *gapDH-C* | 143 | 64 | F: CGC GGC AGA AAA TAT CAT TCC C<br>R: GAA TCG ATA CCA GTT CAG TGA CC |
| papC_1* | *papC* | 109 | 64 | F: TTT CAT GGG TTG CCG GGA GTG<br>R: AGC ATC AGT CCG GGA ATG CC |
| papC_2* | *papC* | 152 | 64 | F: GGG AGT GGA TAT TCG TCC TGA<br>R: ACT GAA CTG ATG AGA GTC TCC TC |
| papGII_1* | *papGII* | 142 | 64 | F: ATT GTC TTT TAC TCC CTT GG<br>R: CCA CGC AAT ATA CTC CCT |
| papGII_2* | *papGII* | 142 | 59/64 | F: TCA TTT CGC GAG TTA CTT GG<br>R: GCC GCA TAA TGA TTA TTA GCG |
| Ia2-383f /ia2-572r | *papGII* | 190 | 64 | F: GGG ATG AGC GGG CCT TTG AT<br>R: CGG GCC CCC AAG TAA CTC G |
| prs-198f/ prs-455r | *papGIII* | 214 | 64 | F: GGC CTG CAA TGG ATT TAC CTG G<br>R: GGC CTG CAA TGG ATT TAC CTG G |
| uidA405-F/uidA-R | *uidA* | 230 | 59/64 | F: CAA CGA ACT GAA CTG GCA GA<br>R: CAT TAC GCT GCG ATG GAT |

**Table 2** summarises the composition of the endpoint PCR reactions and **Table 3** the thermocycler program used. The concentration of the genomic DNA was determined using Qubit (Invitrogen, Waltham, USA) and ranged between 21ng/µl - 114ng/µl.

**Table 2:** Composition of the Endpoint PCR reactions

| Component | Volume [µl] | Finale Concentrations |
|---|---|---|
| 5x Reaction Buffer | 5.0 | 1X |
| dNTPs [10 mM] | 0.5 | 200 µM |
| Q5 High-Fidelity DNA-Polymerase | 0.25 | 0.02 U/µL |
| Forward Primer [10µM] | 1.25 | 0.5 µM |
| Reverse Primer [10µM] | 1.25 | 0.5 µM |
| Template DNA | 2 | variable |
| Nuclease-free Water | 14.75 | |
| Total | 25 | |

**Table 3:** Thermocycler program used for the Endpoint PCR reaction.

| Process | Temperature [°C] | Time [seconds] | Cycles |
|---|---|---|---|
| Initial denaturation | 98 | 30 | 1 |
| Denaturation | 98 | 10 | 32 |
| Primer annealing | 64/59 | 15 | 32 |
| Elongation | 72 | 120 | 1 |
| Finale extension | 72 | 120 | 1 |
| Hold | 12 | ∞ | 1 |

Using the endpoint PCR design summarised in **Table 4**, we assessed the functionality of the primers and tested for cross-reactions between the primers designed for *papGII* and *papGIII*, as there are the two *papG* variants which are genetically most similar (3).

**Table 4:** Endpoint PCR experimental design to test for the functionality of all primers and the cross reactivity between the primers for *papGII* and *papGIII*.

| Name | Target | ***E. coli*** **strain** | ***papG*** **variant** | Expected Outcome |
|---|---|---|---|---|
| gapC_1 | *gapDH-C* | 113269-19 | no papG | + |
| gapC_2 | *gapDH-C* | 113269-19 | no papG | + |

| | | | | |
|---|---|---|---|---|
| papC_1 | *papC* | 700223-20 | papGII | + |
| papC_2 | *papC* | 700223-20 | papGII | + |
| papGII_1 | *papGII* | 113269-19 | papGII | + |
| papGII_2 | *papGII* | 113269-19 | papGII | + |
| Ia2-383f /ia2-572r | *papGII* | 113269-19 | papGII | + |
| prs-198f/prs-455r | *papGIII* | 711763-19 | papGIII | + |
| uidA405-F/uidA-R | *uidA* | 113269-19 | no papG | + |
| gapC_1 | *gapDH-C* | 145484-19 | papGII | + |
| gapC_2 | *gapDH-C* | 145484-19 | papGII | + |
| papC_1 | *papC* | 113269-19 | no papG | - |
| papC_2 | *papC* | 113269-19 | no papG | - |
| papGII_1 | *papGII* | 711763-19 | papGIII | - |
| papGII_2 | *papGII* | 711763-19 | papGIII | - |
| Ia2-383f /ia2-572r | *papGII* | 711763-19 | papGIII | - |
| prs-198f/prs-455r | *papGIII* | 145484-19 | papGII | - |
| uidA405-F/uidA-R | *uidA* | 711763-19 | papGII | + |
| papC_1 | *papC* | 700223-20 | papGII | + |

The resulting PCR products were separated using a 2% agarose gel electrophoresis in 1X TAE buffer solution, at 40 Volt during 50 minutes. We used the ‘Safe Red’ dye to visualise the products and recorded the gel on a ‘Gel Visualizer Fusion X’.

##### Centre 2

At centre 2, two sets of primers and probes for the *E. coli* core gene *rpoD* were newly designed using Primer-BLAST (4) (**Table 5**). Both sets were tested for their functionality using endpoint PCR. We used 1 µl of *E. coli* frozen stocks (*E. coli* strains isolated in routine diagnostics and preserved in 1ml of skim milk media) as template.

**Table 5:** Primers which were used in the endpoint PCR assay at centre 2. Primers with an asterisk (*) were newly designed for this study.

| Name | Target | Temp. [°C] | Sequence (5‘-3‘) |
|---|---|---|---|

| | | | |
|---|---|---|---|
| rpoD_1* | *rpoD* | 59<br><br>59.5 | F: GAC GAA GAT GCT GCC GAA G<br>R: CAA TTT CGC CTT CGC GGG<br>Probe: TTC AAC GGT GCC CAT TTC AC |
| rpoD_2* | *rpoD* | 59<br><br>58.5 | F: AGA TGC TGC CGA AGC CG<br>R: TAG CGA TGT CAA TTT CGC CTT<br>Probe:ATG TAC ATG CGT ACC GGG TC |

#### Quantitative PCR

##### Centre 1

In the next step, we evaluated the performance of our primers in a quantitative PCR (qPCR) assay. **Table 6** summarises the composition of the qPCR reactions and **Table 7** summarises the thermocycler program used.

**Table 6:** Composition of the qPCR assay performed at centre 1

| Component | Volume [µl] | Finale Concentrations |
|---|---|---|
| Luna Universal Probe qPCR Master Mix* | 10 | 1X |
| Probe [10µM] | 0.4 | 0.2 µM |
| Forward Primer [10µM] | 0.8 | 0.4 µM |
| Reverse Primer [10µM] | 0.8 | 0.4 µM |
| Template DNA | 1 | variable |
| Nuclease-free Water | 7 | |
| Total | 20 | |

**Table 7:** Thermocycler program used for the qPCR reaction performed at centre 1

| Process | Temperature [°C] | Time [seconds] | Cycles |
|---|---|---|---|
| Initial denaturation | 95 | 60 | 1 |
| Denaturation | 95 | 15 | 40 |
| Primer annealing | 64/59 | 15 | 40 |
| Extension | 60 | 30 | 40 |

We tested the specificity of the primers gapC_2, uidA, papC_1 and papGII on 32 strains (**Table 8**).

**Table 8:** 32 strains used to assess the specificity of the selected PCR primers uidA, gapC_2, papC_1 and papGII_2. 'ST' = Sequence Type.

| Strain | Phylogroup | ST | H-ype | O-type | Capsule Type | papG variant |
|---|---|---|---|---|---|---|
| 113269-19 | A | 1972 | H30 | O147 | no capsule | no papG |
| 103713-49 | B1 | 58 | H25 | O9 | no capsule | no papG |
| 100223-19 | B2 | 73 | H1 | O25 | KX21 | no papG |
| 128632-18 | C | 1426 | H16 | O8 | no capsule | no papG |
| 100080-19 | D | 362 | H31 | O23 | KX75 | no papG |
| 712184-19 | E | 1011 | H45 | O166 | no capsule | no papG |
| 117680-19 | F | 3058 | H24 | O8,O112 | no capsule | no papG |
| 702833-20 | G | 1163 | H23 | O171 | no capsule | no papG |
| 145484-19 | B2 | 131 | H5 | O16 | KX05 | papGII |
| 700223-20 | B2 | 131 | H4 | O25 | KX21 | papGII |
| 106880-19 | B2 | 131 | H4 | O25 | KX41 | papGII |
| 107962-20 | B2 | 131 | H4 | O25 | no capsule | papGII |
| 100033-19 | B2 | 95 | H7 | O1 | KX03 | papGII |
| 100888-20 | B2 | 73 | H1 | O6 | KX29 | papGII |
| 123488-18 | B2 | 73 | H1 | O6 | KX41 | papGII |

| | | | | | | |
|---|---|---|---|---|---|---|
| 711667-19 | B2 | 73 | H1 | O6 | no capsule | papGII |
| 127688-19 | D | 69 | H18 | O15 | K96 | papGII |
| 109841-19 | D | 69 | H4 | O117-Gp8 | KX31 | papGII |
| 721814-18 | F | 62 | H45 | O7 | KX03 | papGII |
| 135695-19 | F | 648 | H2 | O153var1 | KX42 | papGII |
| 711763-19 | A | 93 | H4 | O5 | KX21 | papGIII |
| 130292-18 | B1 | 75 | H8 | O112 | no capsule | papGIII |
| 118504-18 | B2 | 131 | H4 | O25 | KX21 | papGIII |
| 102689-19 | B2 | 12 | H5 | O4 | KX53 | papGIII |
| 700699-20 | C | 88 | H9 | O8 | no capsule | papGIII |
| 116138-19 | G | 117 | H4 | O143 | no capsule | papGIII |
| 103840-19 | D | 69 | H1 | O15 | KX47 | papGIV |
| 701188-20 | D | 38 | H30 | O153var1 | KX21 | papGIV |
| 716875-19 | F | 59 | H7 | O1 | KX42 | papGIV |
| 720707-18 | B2 | 73 | H1 | O6 | KX24 | papGV |
| 102916-19 | C | 367 | H19 | O9 | no capsule | papGV |
| 720603-19 | D | 69 | H18 | O15 | KX02 | papGV |

##### Centre 2

We compared the efficiency of the two newly designed *rpoD* probes in a qPCR assay, using the composition summarised in **Table 9** and the thermocycler program summarised in **Table 10.**

**Table 9:** Composition of the qPCR assay performed at centre 2

| Component | Volume [µl] | Finale Concentrations |
|---|---|---|
| Luna Universal Probe qPCR Master Mix* | 10 | 0.48X |
| Probe [2.5µM] | 1 | 0.12 µM |
| Forward Primer [20µM] | 0.5 | 0.48 µM |
| Reverse Primer [20µM] | 0.5 | 0.48 µM |
| Template (*E. coli* frozen stocks) | 1 | variable |
| Nuclease-free Water | 8 | |
| Total | 21 | |

**Table 10:** Thermocycler program used for the qPCR reaction performed at centre 2

| Process | Temperature [°C] | Time [seconds] | Cycles |
|---|---|---|---|
| Initial denaturation | 95 | 300 | 1 |
| Denaturation | 95 | 15 | 40 |
| Primer annealing | 60 | 10 | 40 |
| Extension | 60 | 30 | 40 |

#### Multiplexing the qPCR

##### Centre 1

To further decrease the turnaround time of our assay, we further assessed the possibility to combine our primer to a multiplex assay (composition summarised in **Table 11**).

**Table 11:** Composition of the qPCR assay, adapted for multiplexing used at centre 1

| Component | Volume [µl] | Finale Concentrations |
|---|---|---|

| | | |
|---|---|---|
| Luna Universal Probe qPCR Master Mix* | 10 | 1X |
| Forward Primer 1 [10µM] | 0.8 | 0.4 µM |
| Reverse Primer 1 [10µM] | 0.8 | 0.4 µM |
| Forward Primer 2 [10µM] | 0.8 | 0.4 µM |
| Reverse Primer 2 [10µM] | 0.8 | 0.4 µM |
| Probe 1 [10µM] | 0.4 | 0.2 µM |
| Probe 2 [10µM] | 0.4 | 0.2 µM |
| Template DNA | 1 | variable |
| Nuclease-free Water | 5 | |
| Total | 20 | |

##### Centre 2

At centre 2, the multiplex assays were performed as depicted in **Table 12**.

**Table 12:** Composition of the qPCR assay, adapted for multiplexing used at centre 2

| **Component** | **Volume [µl]** | **Finale Concentrations** |
|---|---|---|
| Luna Universal Probe qPCR Master Mix* | 10 | 0.48X |
| Probe 1 [2.5µM] | 1 | 0.12 µM |
| Forward Primer 1 [20µM] | 0.5 | 0.48 µM |
| Reverse Primer 1 [20µM] | 0.5 | 0.48 µM |
| Probe 2 [2.5µM] | 1 | 0.12 µM |
| Forward Primer 2 [20µM] | 0.5 | 0.48 µM |
| Reverse Primer 2 [20µM] | 0.5 | 0.48 µM |
| Template (*E. coli* frozen stocks) | 1 | variable |
| Nuclease-free Water | 6 | |
| Total | 21 | |

#### Applying qPCR assay directly to urine samples

##### Centre 1

Urine samples were stored at 4°C and were pelleted as previously described (52). Urine samples (1 ml) were centrifuged (10 minutes, 5,000 rounds per minute (rpm)), the supernatants were removed and the pellets were resuspended in 1ml nuclease free water. Samples were centrifuged again (5 minutes, 10,000 rpm), the supernatants were removed and the resulting pellets were resuspended in 50 µl nuclease free water. Samples were then heated to 99°C on a Thermomixer heat block (Eppendorf, Hamburg, Germany) for 10 min. We assessed the efficiency and the limit of detection our qPCR assay performed on samples processed using this method, and compared them to values resulting from qPCR on extracted genomic DNA.

#### Screening of patient samples

##### Centre 1

We prospectively collected 543 urine samples, which we processed to urine pellets before boiling (see methods section). All samples were screened for *gapC* and *papC* in a duplex qPCR approach (see composition in **Table 11** and cycling parameters in **Table 7**). In a second experiment, we screened samples which were culture positive for *E. coli* or for which *gapC* had been detected in the multiplex qPCR for the presence of *papGII* using a single qPCR (see composition in **Table 6** and cycling parameters in **Table 7**). 150 multiplex and 74 single *papGII* reactions were performed in duplicate, the remaining ones as single measurements. Samples for which the duplicate measurements had a difference of > 1.5 Ct values in any of the three targets were excluded from further analysis. For the single PCR of *papGII* we further set a cut-off to 36 cycles. Only samples for which a signal was recorded in less than 36 cycles were considered *papGII* positive. All *papGII* positive measurements were furthermore manually curated and measurements for which the amplifications curve was discontinued were set to negative.

##### Centre 2

We prospectively collected 1,106 *E. coli* strains, for which an AMR profile was measured in routine diagnostics. Strains were either isolated from urine (n=1,011) or from blood culture (n=95) samples of 886 patients. We used 10µl plastic inoculation needles to transfer bacterial material from Müller-Hinton agar plates to 1ml of skim milk freezing media and stored all strains at -80°C. Frozen stocks were thawed at room temperature before being subjected to qPCR. All samples were initially screened for *rpoD* and *papC* (primers and probes tested at centre 1). Samples which tested positive for *papC* were further subjected to a duplex qPCR amplifying *rpoD* and *papGII* (see composition in **Table 12** and cycling parameters in **Table 10**). All samples and targets were manually reviewed, using a Ct-value of 25 as threshold and setting samples with discontinued amplification curves to negative. Measurements which were tested negative for *rpoD* were excluded from further analyses.
