## Supplementary material for "Bacterial genome wide association study substantiates *papGII* of *Escherichia coli* as a patient independent driver of urosepsis": Supplementary PCR Data.pdf

### Evaluation of primer functionality

We tested the functionality of eleven primer pairs using an endpoint PCR. For all primer pairs except for prs-198f/prs-455r the expected signal could be recorded and no cross reactivity between the *papGII* primers and *papGIII* target was detected (**Figure S14**). prs-198f/prs-455r was excluded from further analysis. In a next step, we evaluated the limit of detection and efficiency of the remaining primers in a qPCR assay (**Table S3**, **Figure S15**) and tested the specificity of the primers gapC_2, uidA, papC_1, and papGII_2 using 32 *E. coli* strains of which 24 carried a *pap* operon and 12 the *papG* variant *papGII*. An amplification was recorded in all expected reactions (100/100) and no unexpected signal was recorded using the *papC* primers (8/8) nor the papGII_2 primers (20/20). Based on the recorded values we selected the primers gapC_2, papC_1, and papGII_2 for further analysis at centre 1 and rpoD_2, papC_1, and papGII_2 at centre 2 (**Table S4**).

**Table S4:** Primer pairs designed for and included in this study. 'For.'= Forward primer; 'Rev.'= Reverse primer.

| Primer | Nucleotide sequence | Annealing temperature | Used at |
|---|---|---|---|
| gapC_2 | For.: CGC GGC AGA AAA TAT CAT TCC C<br>Rev.: GAA TCG ATA CCA GTT CAG TGA CC<br>Probe: ACG CGT GCC GGT GAA AAC AGG | 64°C | Centre 1 |
| rpoD_1 | For.: GAC GAA GAT GCT GCC GAA G<br>Rev.: CAA TTT CGC CTT CGC GGG<br>Probe: ATG TAC ATG CGT ACC GGG TC | 60°C | Centre 2 |
| papC_1 | For.: TTT CAT GGG TTG CCG GGA GTG<br>Rev.: AGC ATC AGT CCG GGA ATG CC<br>Probe:TGA TGC CAC CTG GCT GCC TCC CT | 64°C | Centre 1 & 2 |
| papGII_2 | For.: TCA TTT CGC GAG TTA CTT GG<br>Rev.: GCC GCA TAA TGA TTA TTA GCG<br>Probe: AAG AAT ATC GGC GGA TGC CGT C | 64°C | Centre 1 & 2 |

We examined whether variants of these sequences occur in our set of genomes (n=1,076). In >90% of sequences in which all sites of the target had a coverage exceeding the threshold from the variant caller, an exact match of the primer and probe sequences was detected (**Figure S16**). We further assessed the possibility of multiplexing our primers: *gapC*, *rpoD* and *papC* were reliably amplified in duplex reactions (gapC_2/papC_1, rpoD_1/papGII_2) as well as in triplex reactions (gapC_2/papC_1/papGII_2 or rpoD/papC_1/papGII_2) (**Figure S17**). In contrast, whole papGII was reliably amplified in duplex reactions (rpoD_1/papGII_2), it was not reliably amplified in a triplex reaction (neither gapC_2/papC_1/papGII_2 nor rpoD/papC_1/papGII_2) (**Figure S17**). We therefore used gapC_2 and papC_1 in a duplex reaction and screened for *papGII* either in a single reaction (centre 1) or in a duplex reaction amplifying *rpoD* (centre 2). We aimed to apply our assay directly to pelleted urine samples, thereby omitting overnight culturing. We observe a 10-fold increase in the limit of detection for papC_1 and papGII_2 (**Table S3**) doing so compared to when using extracted DNA. When comparing pelleted urine samples (n=24) which were boiled to those whose DNA was extracted using the QIAamp DNA Mini Kit, we observe a high correlation between the resulting Ct-values (**Figure S18**). Only for the papC_1 primer, however, a signal was detected for extracted DNA but not for boiled urine pellets in 11/24 samples.
